# The role of the metabolic profile in mediating the relationship between body mass index and left ventricular mass in adolescents: analysis of a prospective cohort study

**DOI:** 10.1101/2020.03.09.20033324

**Authors:** Alice R Carter, Diana L. Santos Ferreira, Amy E Taylor, Deborah A Lawlor, George Davey Smith, Naveed Sattar, Nishi Chaturvedi, Alun D Hughes, Laura D Howe

**Affiliations:** MRC Integrative Epidemiology Unit, Population Health Sciences, University of Bristol, Bristol, UK; National Institute for Health Research Biomedical Research Centre at the University Hospitals Bristol NHS Foundation Trust and the University of Bristol, Bristol, UK; Institute of Cardiovascular and Medical Science, University of Glasgow, Glasgow, UK; Institute of Cardiovascular Science, University College London, London, UK

**Keywords:** metabolic profile, adiposity, mediation, cardiac structure, ALSPAC

## Abstract

**Background:** We aimed to quantify the role of the plasma metabolic profile in explaining the effect of adiposity on cardiac structure.

**Methods:** Body mass index (BMI) was measured at age 11 in the Avon Longitudinal Study of Parents and Children. Left ventricular mass indexed to height^2.7^ (LVMI), was assessed by echocardiography at age 17. The metabolic profile was quantified via nuclear magnetic resonance spectroscopy at age 15.

Multivariable confounder (maternal age, parity, highest qualification, maternal smoking, pre-pregnancy BMI, pre-pregnancy height, household social class and adolescent birthweight, adolescent smoking, fruit and vegetable consumption, physical activity) -adjusted linear regression estimated the association of BMI with LVMI and mediation by metabolic traits. We considered 156 metabolomic traits individually, jointly as principal components (PCs) explaining 95% of the variance in the NMR platform, and assessed whether the PCs for the metabolic traits added to the proportion of the association explained by established cardiovascular risk factors (systolic and diastolic blood pressure, insulin, triglycerides, low density lipoprotein, and glucose).

**Results:** A 1kg/m^2^ higher BMI was associated with a 0.70 g/m^2.7^ (0.53, 0.88) and 0.66 g/m^2.7^ (0.53, 0.79) higher LVMI in males (N=437) and females (N=536), respectively. Established risk factors explained 3% (95% CI: 2% to 5%) of this association in males, increasing to 10% (95% CI: 8%, 13%) when including metabolic PCs. In females, the standard risk factors explained 3% (95% CI: 2%, 5%) of the association, and did not increase when including the metabolic PCs.

**Conclusion:** The addition of the NMR measured metabolic traits appear to mediate more of the effect of BMI on LVMI than the established risk factors alone in adolescent males, but not females.

## Introduction

Cardiovascular disease (CVD) remains the leading cause of death globally (1), and adiposity is a key CVD risk factor (2). Mediation analysis can be used to gain a wider aetiological understanding of an exposure, in addition to identifying modifiable intermediate variables linking the exposure to a particular outcome (3). Interventions to prevent or treat high levels of adiposity have had limited impact, therefore, identifying novel modifiable intermediate processes between adiposity and CVD provide an opportunity for future interventions aiming to reduce risk of CVD (4-6).

Blood pressure, glucose, insulin and lipid levels have been identified as major contributors to the association between adiposity and CVD. These factors have been estimated to explain 46% and 76% of the association between BMI and coronary heart disease and stroke respectively (7). The availability of metabolomic data in cohort studies, specifically the numerous lipid-based measures determined via nuclear magnetic resonance (NMR) spectroscopy, has led to an increased understanding of the causal effects of body mass index (BMI) on circulating metabolites (8) as well as the role of such metabolites on CVD risk (9, 10). Therefore, metabolic intermediates are strong candidates as intermediates on the causal pathway from adiposity to CVD risk, which importantly, can be intervened on. For example, harmful cholesterol levels are already targeted using statin medication, which is widely prescribed in routine general practice.

Although adverse cardiovascular events largely occur in adult life, cardiovascular pathology has been shown to have its origins in early life (11-14), with levels of adiposity and cardiovascular risk factors known to track from childhood through to adulthood (15). Measures of cardiac structure and function in adults are preclinical markers of CVD (16), and there is evidence that cardiac structure in young adults is associated with future risk of CVD events (17). Previous analyses carried out in the cohort used in this study (the Avon Longitudinal Study of Parents and Children (ALSPAC)) have demonstrated a causal relationship between body mass index (BMI) and left ventricular mass indexed to height^2.7^ (LVMI), a measure of cardiac structure, in adolescents (18).

In this study, we use data from adolescents in ALSPAC, a UK prospective cohort study, to assess the role of NMR-measured metabolic traits as mediators of the association between BMI and LVMI. Mediation analysis is inherently a causal inference method, where causality is assumed between the exposure and outcome, exposure and mediator and mediator and outcome (3). Therefore, these analyses focus on the association between BMI and LVMI given the existing evidence for a causal relationship (18). Our primary aim is to identify whether considering the whole of the NMR-measured metabolic profile results in a greater proportion of the BMI-LVMI relationship being explained over and above the amount explained by established risk factors (systolic blood pressure [SBP], diastolic blood pressure [DBP], insulin, triglycerides, low density lipoprotein cholesterol [LDL-C], and glucose).

## Methods

### Participants

ALSPAC is a population-based birth cohort study. Pregnant women living in the former county of Avon, South West England, with an expected delivery date between 1 April 1991 and 31 December 1992 were eligible for enrolment. In total, 14,541 women were enrolled in to ALSPAC, with 14,901 children born. The participants have been followed up since birth, with questionnaires, links with routine data and research clinics. Full details of the cohort have been reported previously (19, 20). Ethical approval was obtained from the ALSPAC Law and Ethics committee and local ethics committees. The study website contains details of all the data that is available through a fully searchable data dictionary and variable search tool (http://www.bristol.ac.uk/alspac/researchers/our-data/) (21). To maintain temporal sequencing of our exposures, mediators and outcomes, we used adiposity measures from age 11, metabolic traits assessed at age 15, and cardiac structure assessed at age 17 years.

### Anthropometric measurements

At the age 11 follow up clinic height was measured using the Harpenden Stadiometer, without shoes. Weight was measured using the Tanita Body Fat Analyser. BMI was then calculated as weight in kg divided by the square of height in metres.

### Mediator measurements

Fasting (overnight or minimum 6-hours) plasma metabolic traits were quantified via high-throughput ^1^H-Nuclear magnetic resonance spectroscopy (NMR), Nightingale Health© (Helsinki, Finland), at age 15. For samples taken in the morning the fasting period was overnight and for afternoon samples (after 14:00) individuals were required to fast for at least 6 hours. The 156 metabolic traits included in analyses span various metabolic pathways (mostly lipid and lipoprotein related) (Supplementary Table 1).

Established mediators were measured using fasting plasma glucose samples. Fasting plasma glucose was measured using an automated assay. Insulin was measured from blood samples using an enzyme-linked immunosorbent assay (Mercodia, Uppsala, Sweden). Plasma lipid concentrations, including triglycerides and LDL-C, were taken from venous blood samples and measured by using enzymatic reagents for lipid determination. The Friedewald equation was used to estimate LDL-C (22). Where traits, such as LDL-C are measured in both the NMR platform and as established mediators from plasma glucose, the traits were excluded from the NMR platform (see statistical analysis section for full details).

Resting systolic blood pressure and diastolic blood pressure were measured at least twice during clinics, using a Dinamap 9301 Vital Signs Monitor (Morton Medical, London) and cuff size appropriate for the child. A mean of the final two measures was used.

### Cardiac structure measures

Left ventricular mass was assessed by echocardiography in a quasi-random subset of participants in ALSPAC at the age 17 clinic. Echocardiography was performed using a HDI 5000 ultrasound machine (Philips) equipped with a P4-2 phased-array ultrasound transducer. All measurements were made according to the American Society of Echocardiography guidelines, and validated equations were used to calculate left ventricular mass, which was indexed to height^2.7^ (LVMI) (23).

### Confounder assessment

Mediation assumes causal effects and therefore that there is no confounding between the exposure and outcome, exposure and mediator and mediator and outcome, as well as no intermediate confounders (that being a confounder of the mediator and outcome that is itself influenced by the exposure) (3). Confounders included in analyses were selected based on *a priori* knowledge and were included in all analysis models as either confounders of the exposure and mediator, mediator and outcome, exposure and outcome or between all three (see Figure 1). Maternal confounders in this analysis were age, parity, education, pre-pregnancy height, pre-pregnancy BMI, and smoking. Adolescent confounders were birthweight, smoking (at age 15), physical activity (at age 15) and diet (at age 15) measured by fruit and vegetable intake. Household social class, around the time of pregnancy, was also included as a confounder. Full details of all confounders and their measurement are in the Supplementary Material.

**Figure 1:**
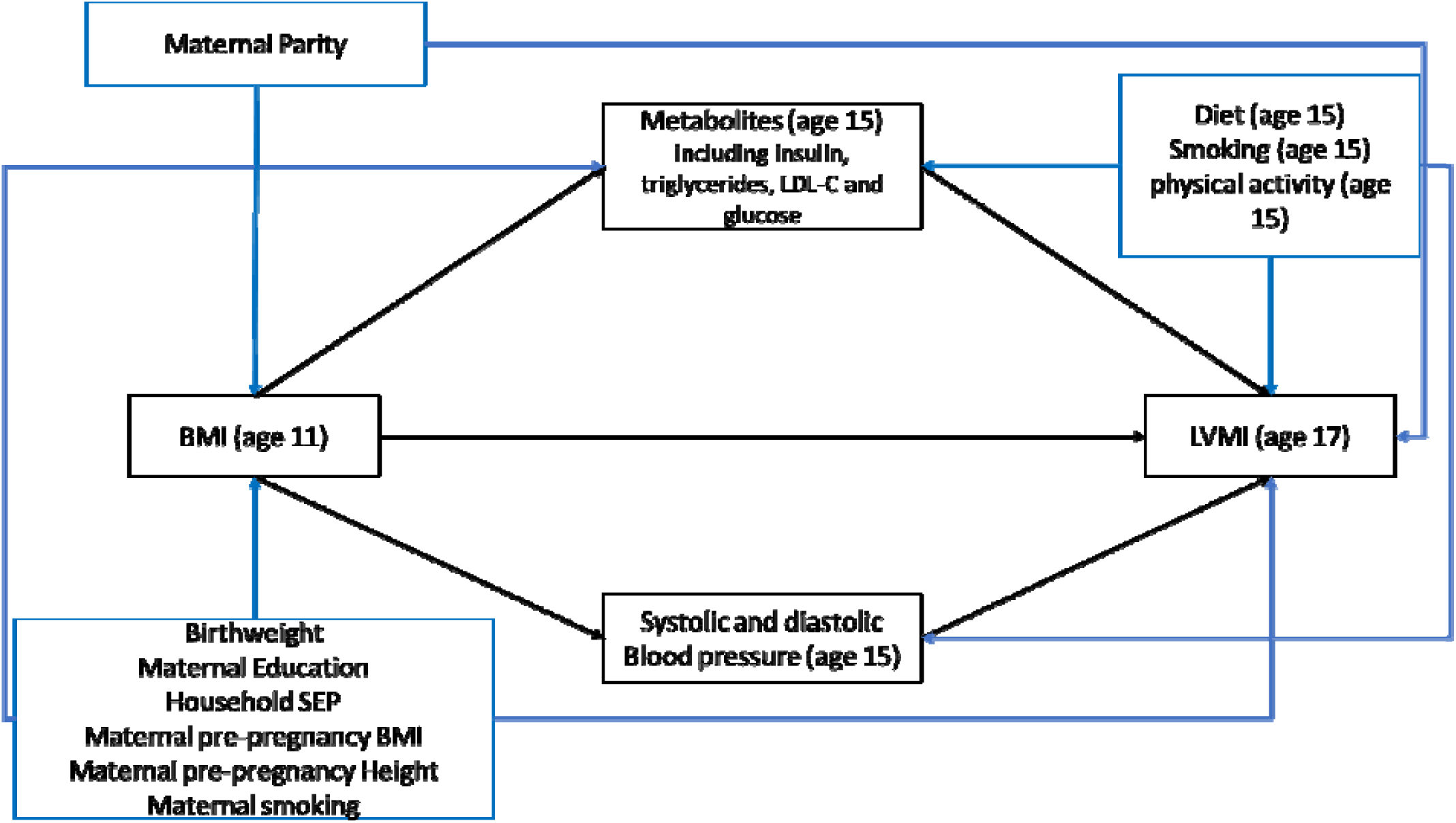
Directed acyclic graph depicting causal assumptions made in mediation analyses assessing the role of metabolic mediators on the association between body mass index and left ventricular mass indexed to height^2.7^.

Participants were excluded if a value below zero was recorded for any anthropometric trait (BMI, waist circumference and DXA-determined fat mass) (N=22 excluded). Additionally, one individual was excluded as they were an analytical outlier on the NMR platform. Confounders with a value below zero (mainly reflecting missing data) were recoded as missing and multiply imputed as with other missing data (Supplementary Table 2).

### Statistical analysis

All analyses were run on Stata 15. Based on previous literature indicating different cardiac risk profiles in males and females, it was decided *a priori* to carry out all analyses stratified by sex (24-27).

Multivariable linear regression was used to test the association between i) BMI and LVMI (total effect ii) the association between BMI and each metabolic trait individually and iii), the association between each individual metabolic trait and LVMI. All analyses were adjusted for the confounders specified in the previous section.

As mediation analyses assumes causal effects it uses a terminology (e.g. total effects) to reflect that, as we do here (we discuss the extent to which the assumptions of mediation analyses are likely to be violated in the discussion section of the paper). Several mediation models were carried out to assess the extent to which the total effect was explained by the metabolomic profile and established risk factors. The models considered were i) each metabolic trait considered individually, ii) all traits in the NMR metabolic platform considered together (as principal components), iii) a set of ‘established’ cardiovascular risk factors (SBP, DBP, insulin, triglycerides, LDL-C, and glucose), iv) the established cardiovascular risk factors and NMR metabolic traits (as principal components, described below) together. When considered individually, the NMR metabolic traits were standardized to set the means to 0 and standard deviations to 1.

Mediation was assessed in a counterfactual framework, where interactions between BMI and NMR metabolic traits were allowed in individual mediation models, and multiple mediator models we assumed no interaction between BMI and the mediators (28). We report natural direct effects (the effect of BMI on LVMI not via mediators, for a one kg/m^2^ increase in BMI where the value of the mediator is allowed to vary for each individual) and natural indirect effects (the mediated effect of the association between BMI and LVMI, for a one SD increase in NMR metabolic traits) (28, 29) (3). The confidence interval for the indirect effect was obtained via bootstrapping with 1000 replications. The proportion mediated is calculated by dividing the indirect effect by the total effect and confidence intervals derived by bootstrapping.

### Principal components for metabolic traits

In multiple mediator analyses considering multiple NMR metabolic traits in a single model (models ii to iv), principal components (PCs) of the standardized values of the NMR metabolic traits were used to account for collinearity. The inclusion of multiple collinear variables in a model can result in inflated standard errors (30).

Principal component analysis is a data reduction technique, taking a set of correlated variables and extracting a set of uncorrelated PCs. Each PC is a linear combination of the original variables in the data (31).

A number of established risk factors (insulin, triglycerides, LDL-C, and glucose) are included in the NMR metabolic traits. To avoid double counting these mediators in models considering the role of the NMR metabolic traits in addition to established risk factors (model iv), the NMR measurements of these established risk factors were excluded when generating the PCs.

PCs were estimated separately for males and females. For use in mediation analysis, we included the number of PCs required to estimate 95% of the variance in the NMR metabolic traits. For model ii (all NMR metabolic traits), this was 18 PCs in the females and 19 PCs in the males. For model iv (established risk factors plus NMR metabolic measures), 20 PCs were included in the analysis of females and 21 PCs for males. Taken together, these PCs capture variation across the metabolic profile. Therefore, we cannot use these analyses to identify the contribution of specific metabolic traits to mediation.

### Multiple Imputation

To maximise power and potentially reduce bias, multivariable multiple imputation was carried out to impute missing confounders. The proportion of missingness is available in Supplementary Table 2. The sample for imputation was defined as all individuals with complete data on all adiposity variables at ages 11, mediators (including NMR metabolic platform and established risk factors) at age 15 and echocardiography data at age 17. The PCs reflecting 95% of the variance in all NMR metabolic traits were included in the imputation model, rather than all NMR metabolic traits, to avoid collinearity and convergence problems. We created 20 imputed datasets. The distribution of these imputed variables was assessed to confirm the imputed data was consistent with the original data.

Each imputed dataset was analysed separately, with the results combined using Rubin’s rules.

### Sensitivity analyses

Although sex stratified analyses were pre-specified *a priori*, a likelihood ratio test was carried out to test whether a model for the total effect accounting for interaction by sex was a better fit than when interactions were not considered (24-26).

It was determined *a priori* to use BMI, mediators (including metabolic traits) and LVMI all measured at different timepoints. The pairwise correlation between BMI measures at age 11 and BMI measured at age 15 was assessed to identify whether BMI was stable across puberty.

In addition to BMI, all analyses (including all individual mediator models and all multiple mediator models) were replicated using waist circumference and Dual-X ray absorptiometry [DXA]-determined fat mass as measures of adiposity. Three additional measures of cardiac structure which have been linked to cardiovascular health were also considered in sensitivity analyses, namely, left atrial size indexed to height (LAI), left ventricular internal diameter (LVIDD) and relative wall thickness (RWT). In total, the association between each exposure (BMI, waist circumference and DXA-determined fat mass) was assessed with each outcome (LVMI, LAI, LVIDD and RWT). For each of these exposure and outcome combinations the mediating effect of i) individual metabolic traits ii) PCs for the metabolic profile iii) established risk factors and iv) established risk factors plus PCs for the metabolic profile, was estimated. Full details of the additional adiposity and cardiac structure measurements is available in the Supplementary material.

From the individual metabolic trait mediation results, small very low-density lipoproteins (small VLDL) as a group appeared to have a stronger mediating effect (i.e. a larger indirect effect) than other groups of NMR metabolic traits. Therefore, as a post-hoc sensitivity analysis to understand whether the effects of the NMR metabolic traits considered jointly were driven by the small VLDL class of lipoproteins, we ran sensitivity analyses including only these in a model with established cardiovascular risk factors across all exposure and outcome combinations.

To evaluate whether total effects and indirect effects were independent of puberty, age at peak height velocity (32), an indicator of timing of puberty, was included as a covariate in multiple mediator models assessing mediation between BMI and LVMI with i) the metabolic PCs and ii) joint model with the metabolic PCs and established risk factors.

In addition to analyses using imputed data, complete case analyses were carried out for the association between BMI and LVMI and the extent to which the total effect was explained by the mediators considered in the main analyses.

## Results

### Participant characteristics

A total of 1004 participants were eligible for analysis. Of these, 467 were males and 537 females. A study flow chart is shown in Figure 2.

**Figure 2:**
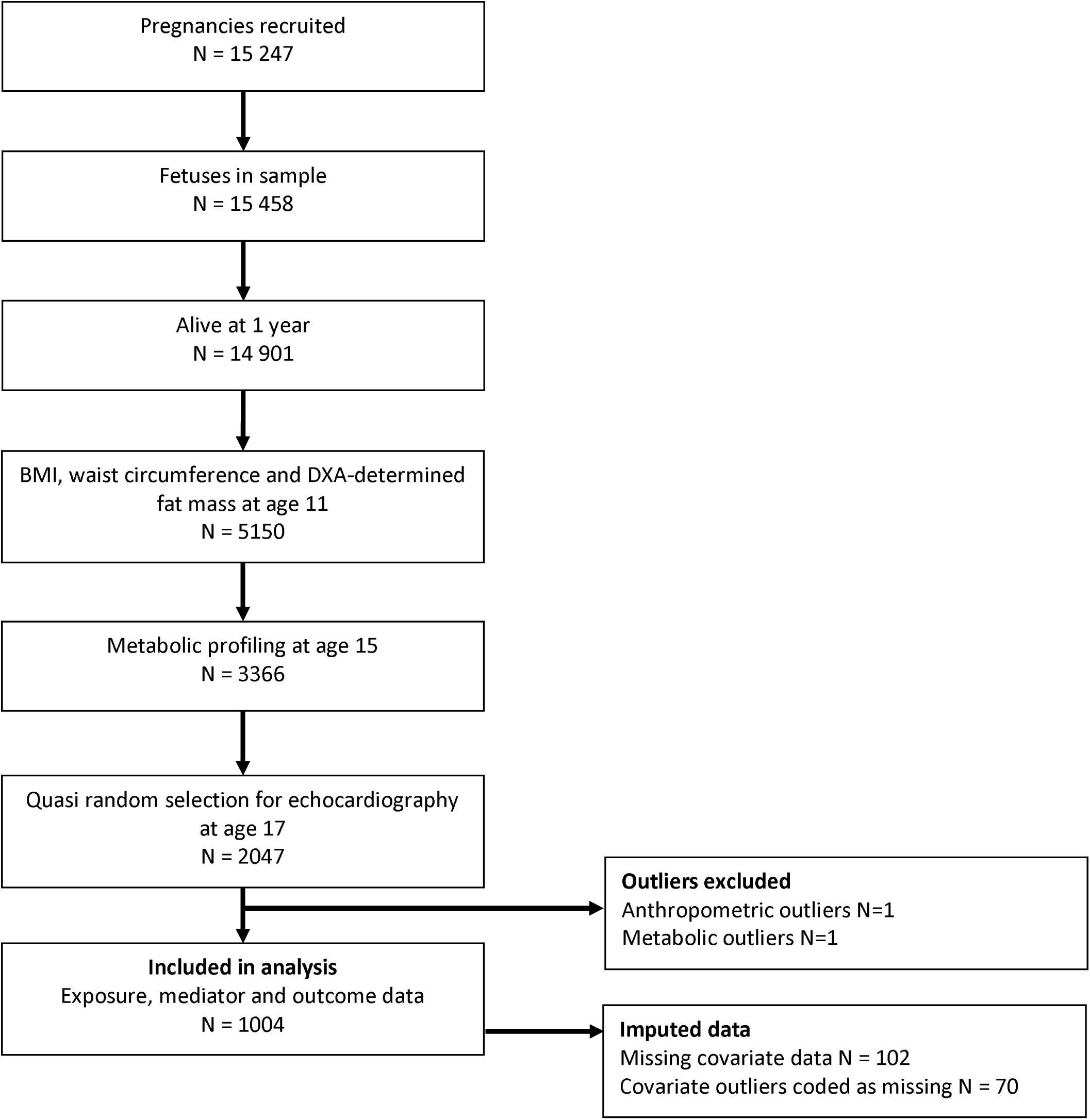
Flow chart of study recruitment to inclusion in analyses. BMI = Body mass index DXA-determined fat mass = Dual X-ray absorptiometry-determined fat mass

Participants included had similar mean BMI, waist circumference and fat mass as all ALSPAC participants who attended the 11-year clinic. Cardiac structure and function measures were also similar between our study sample and all participants who attended the 17-year clinic and had cardiac measures. Participants in the analysis sample were typically from higher social class backgrounds, and there were fewer participants whose mothers had ever smoked and fewer males, compared with the full cohort. The characteristics of the imputed data were comparable with the non-imputed data in the analysis sample. Full participant characteristics are presented in Supplementary Table 3.

### Association between adiposity, risk factors and cardiac structure

A 1 kg/m^2^ higher BMI in females was associated with an increase in mean LVMI of 0.66 g/m^2.7^ (95% CI: 0.53 to 0.79). Similarly, in males, a 1 kg/m^2^ higher BMI was associated with an increase in mean LVMI of 0.70 g/m^2.7^ (95% CI: 0.53 to 0.88) (Table 1). BMI was positively associated with all subclasses of very low-density lipoproteins (VLDL), although the strength of this association was weaker with the very small VLDL subclass. Additionally, the association was typically stronger in males compared with females. There was little evidence of an association between BMI and the low-density lipoproteins (LDL) subclass of metabolic traits. BMI was mostly negatively associated with the high-density lipoprotein (HDL) subclass of metabolic traits There was little evidence of an association between BMI and fatty acids, nor with fatty acid ratios. In both males and females, BMI was negatively associated with citrate. There was evidence of a positive association between BMI and branched-chain amino acids in males, but not in females (Supplementary Figure 1).

**Table 1:** The total effect of body mass index on cardiac structure assessed using multivariable linear regression stratified by sex.

| Exposure (1 kg/m<sup>2</sup> increase) | Outcome | Females mean difference (95% CI) (N=536) | Males mean difference (g/m<sup>2.7</sup>) (95% CI) (N=437) |
| --- | --- | --- | --- |
| **BMI** | *LVMl* | 0.661 (0.529, 0.793) | 0.701 (0.525, 0.877) |
|  | *LAI* | -0.002 (-0.006, 0.001) | -0.006 (-0.016, 0.003) |
|  | *LVIDD* | 0.027 (0.017, 0.036) | 0.012 (0.027, 0.042) |
|  | *RWT* | 0.001 (-0.0001, 0.003) | 0.002 (2.86E-05, 0.004) |

Models adjusted for: Maternal covariables; age, parity, education, pre-pregnancy height, pre-pregnancy BMI, smoking, household social class and adolescent covariables; birthweight, smoking, physical activity and diet

In all VLDL lipoprotein subclasses, metabolic traits were positively associated with LVMI in both males and females. Although the confidence intervals often spanned the null value, particularly in males. With the exception of the triglyceride metabolic traits, large, medium and small LDL traits were positively associated with LVMI. This association was stronger in males, compared with females, where the confidence intervals often spanned the null value. In females, all subclasses of HDL metabolic traits were negatively associated with LVMI. In males, very large and large HDL metabolic traits were negatively associated with LVMI, but this was less consistent with medium and small HDL metabolic traits. There was some evidence in males of a positive association between fatty acids and LVMI, although this was less consistent in females. In both males and females, citrate was negatively associated with LVMI. There was some evidence of an association between branched chain amino acids and LVMI (Supplementary Figure 2).

### Mediation of the association between adiposity and cardiac structure

Considered separately, each metabolic trait explained only a small proportion of the association between BMI and LVMI. In males, the median proportion mediated for the association between BMI and LVMI was 0.5% (95% CI: 0.5% to 0.5%) and the maximum was 9% (95% CI: 9%, 9%) (explained by citrate). In females, the median proportion mediated was 0.3% (95% CI: 0.3% to 0.3%) and the maximum was 3% (95% CI: 3% to 3%) (explained by acetoacetate) (Figure 3).

**Figure 3:**
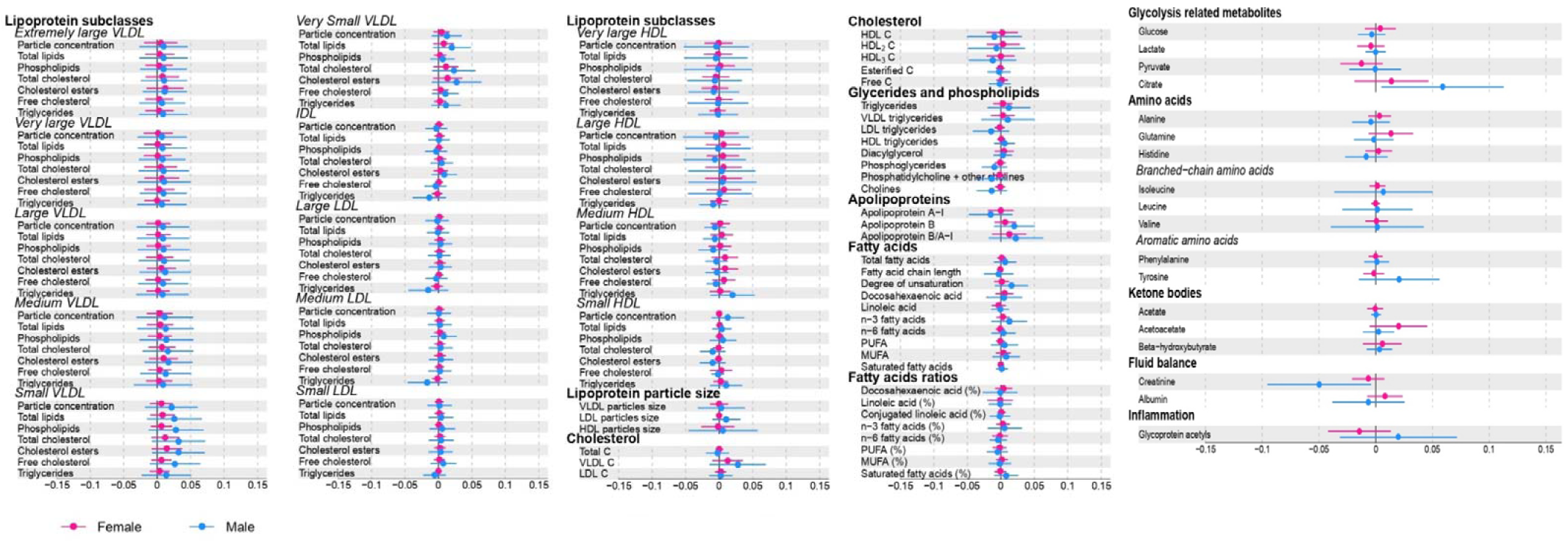
Forest plot showing the natural indirect effect of each NMR metabolic trait individually on the association between body mass index (BMI) to left ventricular mass indexed to height (LVMI) stratified by gender. Models adjusted for: Maternal age, Maternal parity, Maternal education, Maternal pre-pregnancy height, Maternal pre-pregnancy BMI, maternal smoking, household social class and adolescent birthweight, adolescent smoking, adolescent diet and adolescent physical activity VLDL = Very low-density lipoprotein LDL = Low density lipoprotein HDL = High density lipoprotein All results are g/m^2.7^ of LVMI per 1 kg/m^2^ higher BMI

Together, the principal components explaining 95% of variance in the NMR metabolic traits explained 16% (95% CI: 12%, 19%) of the association between BMI and LVMI in males, and 5% (95% CI: 3%, 6%) in females (Table 2).

**Table 2:** Direct and indirect effects of multiple mediator models on the association between body mass index and left ventricular mass indexed to height^2.7^.

| Mediators | Females (N = 536) |  |  | Males (N = 437) |  |  |
| --- | --- | --- | --- | --- | --- | --- |
|  | Natural Direct Effect (95% CI) | Natural Indirect Effect (95% CI) | Proportion Mediated % (95% CI) | Natural Direct Effect (95% CI) | Natural Indirect Effect (95% CI) | Proportion Mediated % (95% CI) |
| **Established risk factors only (systolic blood pressure, diastolic blood pressure, insulin, triglycerides, low density lipoprotein, and glucose)** | 0.64 (0.50, 0.77) | 0.02 (-0.01, 0.06) | 3.46 (1.91, 5.01) | 0.68 (0.50, 0.86) | 0.02 (-0.04, 0.09) | 3.35 (1.72, 4.99) |
| **NMR metabolic measures only (as principal components)** | 0.63 (0.49, 0.77) | 0.03 (-0.03, 0.09) | 4.64 (2.86, 6.41) | 0.59 (0.41, 0.79) | 0.11 (0.003, 0.22) | 15.62 (12.32, 18.91) |
| **Established risk factors + NMR metabolic measures (as PCs)** | 0.65 (0.51, 0.78) | 0.02 (-0.04, 0.06) | 2.34 (1.06, 3.62) | 0.63 (0.44, 0.82) | 0.07 (-0.02, 0.17) | 10.35 (7.59, 13.11) |
| **Established risk factors + Small VLDLs** | 0.63 (0.49, 0.76) | 0.03 (-0.01, 0.07) | 4.81 (3.00, 6.61) | 0.66 (0.48, 0.85) | 0.09 (-0.04, 0.08) | 3.07 (1.51, 4.64) |

Models adjusted for: Maternal covariables; age, parity, education, pre-pregnancy height, pre-pregnancy BMI, smoking, household social class and
adolescent covariables; birthweight, smoking, physical activity and diet. All results are g/m<sup>2,7</sup> of LVMI per 1 kg/m<sup>2</sup> higher BMI

The established cardiovascular risk factors (SBP, DBP, insulin, triglycerides, LDL-C, and glucose) explained 3% (95% CI: 2%, 5%) of the association between BMI and LVMI in males. This increased to 10% (95% CI: 8%, 13%) when the metabolic PCs were included in the model alongside the established risk factors (Table 2).

In females the proportion of the association between BMI and LVMI explained by the established cardiovascular risk factors was 3% (95% CI: 2%, 5%), but when the metabolic PCs were included in the model with the established mediators this reduced to 2% (95% CI: 1%, 4%) (Table 2).

### Sensitivity Analyses

There was little evidence of a statistical interaction between males and females for the total effect of BMI on LVMI (P value_interaction_ = 0.51) (Supplementary Table 4). BMI measured at age 11 was highly correlated with BMI measured at age 15 (pairwise correlation = 0.8), suggesting BMI remains relatively constant during puberty.

The association between waist circumference and separately between DXA-determined fat mass and individual metabolic traits was consistent with the association between BMI and individual metabolic traits. Briefly, in both males and females, both waist circumference and DXA-determined fat mass were positively associated with VLDL subclasses of metabolic traits. There was little evidence of an association between either measure of adiposity and large, medium or small LDL metabolic traits. Consistent with BMI, waist circumference and DXA-determined fat mass were negatively associated with very large, large and medium HDL metabolic traits. As with the association of BMI on metabolic traits, the association between adiposity and individual metabolic traits was typically larger in males compared with females. Estimates of DXA-determined fat mass with metabolic traits were typically estimated more precisely compared with BMI or waist circumference. (Supplementary Figures 3 and 4).

There was little evidence of an association between any individual metabolic trait and LAI or with LVIDD. Although estimates were imprecise, there was greater evidence of an association between individual metabolic traits and RWT. Similar to the association with LVMI, there was a positive trend in the association between VLDL metabolic traits and RWT, and a negative association between large and medium HDL particles (Supplementary Figures 5-7).

In mediation models considering each metabolic trait individually, consistent with the results for the association between BMI and LVMI, each of the NMR metabolic traits explained little of the association between BMI and LAI, LVIDD or RWT. Similarly, each individual metabolic trait mediated little of the association between waist circumference and DXA-determined fat mass with each outcome measure of cardiac structure (Supplementary Figures 8-18).

In multiple mediator analyses, considering BMI as the exposure there was evidence of a stronger mediating effect of the metabolic profile, over and above the amount mediated by established risk factors, for LAI in both males and females. In females, the standard risk factors mediated −6% (95% CI: −8% to −4%) of the association, which increased to 20% (95% CI: 16% to 23 %) when including the metabolic PCs. In males, the established risk factors explained −7% (95% CI: −9% to −5%) of the BMI to LAI association, which increased to 44% (39% to 48%) when the metabolic PCs were included. In females, there was evidence that the metabolic profile mediated more of the effect of BMI on RWT than the established risk factors alone, but not in males.

Considering waist circumference as the exposure, the estimates of mediation in multiple mediator models were similar to those considering BMI as the exposure. There was little evidence in females that the PCs for the metabolic traits mediate more of the effect of waist circumference on LVMI compared with the established risk factors, where 5% of the association was estimated considering both sets of mediators. However, in males, the PCs for the metabolic traits did mediate more of the effect, where the established risk factors mediated 2% (95% CI: 1% to 4%) of the association, which increased to 20% (95% CI: 16% to 24%) when the metabolic PCs were included. This pattern of results was similar when considering DXA-determined fat mass as the exposure. For both waist circumference and DXA-determined fat mass there was greater evidence of mediation by the metabolic traits when considering LAI and RWT as the outcomes, particularly in the females. (Supplementary Figure 19).

In both males and females, there was some evidence of inconsistent mediation by the standard mediators, and in some cases the PCs for the metabolic traits. This inconsistent mediation was most frequently observed when LAI and LVIDD were considered as the outcomes, but there was evidence for inconsistent mediation in all outcomes. Inconsistent mediation occurs when the total effect and indirect effect are in opposing directions leading to a negative proportion mediated. In this case, there is a positive effect of each exposure on the measures of cardiac structure, indicating the indirect effect in these models is negative.

In both males and females, the proportion mediated by total small VLDL were higher than for other metabolic subgroups. In males 4% (95% CI: 4% to 4%) and in females 1% (95% CI: 1% to 1%) of the association between BMI and LVMI was explained by the total lipids small VLDL (Figure 3). Including the small VLDL as mediators for the association between BMI and LVMI made little difference to the estimate of the proportion mediated by established cardiovascular risk factors in either males or females.

In both the males and females, including age at peak height velocity in the models had little effect on the estimates of the proportion mediated (Supplementary Table 5). As such, it is unlikely to be acting as a confounder of the BMI and LVMI association, and more likely to be a mediator of the association between BMI and LVMI. The point estimates for the total effects estimated using complete case data were typically larger than those from multiply imputed data, but with wider levels of imprecision (Supplementary Table 6).

## Discussion

In this cohort of UK adolescents, we have demonstrated that particularly in males, the wider metabolic profile may contribute to the burden of cardiovascular disease attributable to BMI. Including the mainly lipid and lipidomic PCs in addition to established risk factors (SBP, DBP, insulin, triglycerides, LDL-C, and glucose) 10% of the association between BMI and LVMI was explained, compared with 3% by established risk factors alone. In females, the proportion of the association between BMI and LVMI explained by the established risk factors was 3% and was reduced to 2% when including PCs for the metabolic traits. Individually, the metabolic traits explained little of the association between BMI and LVMI. These results were consistent when considering additional measures of adiposity (waist circumference and DXA-determined fat mass) and cardiac structure (LAI, LVIDD and RWT).

Although we see little evidence of strong mediating effects by any single metabolic trait, as a group, the small VLDLs appeared to have a stronger effect than other subgroups. However, we did not find evidence that the small VLDLs are driving the mediating effects of the NMR PCs.

## Results in context

To our knowledge, no other study has looked at the role of NMR metabolic traits as mediators of the association between BMI and LVMI. Using the same data as in this analysis (ALSPAC) a causal effect of BMI and LVMI has been demonstrated (18), providing the motivation for identifying intermediate variables which may mediate this effect. LVMI is a precursor to adverse cardiovascular events in adulthood (33); therefore, identifying intermediate variables from BMI may provide an opportunity to identify potential therapeutic targets.

A recent Mendelian randomization study investigating the mediating effects of lipids and glycaemic traits found stronger mediating effects than our results for the established set of risk factors (34). In our analysis we have considered the role of the metabolic profile in adolescence, whereas in a Mendelian randomization analysis, the estimates reflect a lifetime effect of an exposure (or mediator) (33). Therefore, it may be possible that the mediating role of the metabolic profile between BMI and LVMI (and adiposity and cardiac structure more broadly) emerges throughout the life course.

Sex-differences in cardiometabolic profiles have been shown in a number of studies in both children and adults (25, 26). In a previous study using ALSPAC data, it was shown that the association between BMI and cardiovascular risk factors was stronger in males than females (25). Additionally, sex differences in the association of adiposity and metabolic profile have previously been shown (8). Although we found consistent estimates of the proportion of mediation by the established risk factors in males and females for the association between BMI and LVMI, we found some evidence of stronger mediating effects of the NMR principal components profile in the males. Although there was no strong evidence for a statistical difference between males and females it is likely that we had insufficient statistical power to detect this.

In this analysis, we found less evidence of an association between BMI and individual metabolic traits than previous, larger, analyses. Our smaller sample size is likely to be contributing to these differences (8). Additionally, previous analyses have used Mendelian randomization to explore the causal effect of BMI on individual metabolic traits, which as previously noted will be estimating lifetime effects of an exposure, which may not yet be present in our adolescent cohort.

### Strengths and limitations

In this multivariable regression analysis, residual confounding of associations cannot be ruled out. We controlled for all measured potential confounders of the exposure and outcome, exposure and mediator and mediator and outcome associations, but residual confounding may be present where the variables included in analyses fail to accurately measure the confounder. For example, diet was considered a confounder, and we adjusted for fruit and vegetable intake. However, the confounding effect of diet between BMI and LVMI is likely to be more complex than just considering fruit and vegetable intake. Adolescent smoking was considered as a confounder of the mediator (including metabolic traits and blood pressure traits) and LVMI association in this analysis. However, there is evidence of bidirectional associations between BMI and smoking, where increased BMI is associated with increased smoking (35), and smoking could also be a mediator of BMI and LVMI. As such, there is potential for over adjustment by including smoking in the model. However, in this adolescent cohort we expect the strongest relationship is likely to be smoking influencing the mediators and therefore we adjusted for smoking.

Mediation analysis could be biased by reverse causality due to a mis-specified model, for example if the metabolic profile influenced adiposity rather than the converse. All variables considered were measured prospectively, with appropriate temporal ordering of the exposure, mediators and outcomes, alleviating concerns over reverse causality, or bias from the use of cross-sectional data in mediation analysis (36). Additionally, as an adolescent population, individuals included in these analyses are unlikely to have experienced an adverse major cardiac event or be on preventative medication for cardiovascular diseases (such as statins). This further lessens concerns over reverse causality and potential bias caused by treatment effects.

It is possible that age 11 is too young to clearly identify the effects of BMI on metabolites and subsequently LVMI, particularly as trajectories of BMI are shown to change through puberty (25). However, given the high correlation between BMI at age 11 and BMI at age 15 in this cohort where the pairwise correlation for BMI at both ages was 0.8, the results presented here are unlikely to be biased by trajectories of BMI during puberty.

In addition to reverse causality and residual confounding, mediation analysis can be biased by measurement error, particularly in the mediator (37). This analysis uses objectively measured metabolic data, representing a broad range of metabolic traits, typically not captured by standard biochemical assays. However, these measures are only a snapshot of one time point (age 15) and may not be capturing the full life course effect of these metabolic traits.

Although the primary analyses focussed on the association between BMI and LVMI, other measures of adiposity and cardiac structure were considered in analyses. BMI is often criticised as a poor indicator of overall adiposity, particularly due to its inability to differentiate between lean and fat mass. DXA-determined fat mass may be a better measure for distinguishing between types of body fat and assessing overall adiposity (38). However, consistent with previous analyses in ALSPAC (25) and other cohorts (39), our estimates of mediation were similar when waist circumference of DXA-determined fat mass were considered as exposures instead of BMI.

The ALSPAC sample is a large contemporary cohort with over 14,000 participants enrolled in the original cohort. However, when the analysis was restricted to the subset of individuals with all relevant data on anthropometry, NMR measured metabolic platform, established cardiovascular risk factors and cardiac structure the sample was just over 1000 individuals. Our findings need to be replicated in a larger cohort, particularly if replication could involve using causal inference methods such as Mendelian randomization to triangulate results (40, 41). However, instrumenting the multiple metabolic traits may prove challenging.

A limitation of looking at these mediating effects in a younger cohort, is that some effects of either the exposure or the metabolic profile may only become apparent later in life. As more large-scale biobanks with adult populations release metabolic data, replicating these analyses in adult populations would be important to see whether these results are replicated with clinical CVD events as outcomes.

### Clinical and public health implications

We show that metabolic traits, acting together, mediate some of the effect of BMI on cardiac structure in adolescence. In these analyses, we have not identified a clear intervenable target by a single lipid or metabolic trait, or metabolic group. The PCs included in mediation analysis reflect the variation in metabolic traits across the metabolic profile. To this extent, they are unlikely to be estimating the effect of a single metabolic trait or metabolic group. Rather, they explore the effect across the metabolic profile. Early intervention on these multiple mediators might therefore be a useful strategy to reduce future cardiovascular disease. Future studies examining the effect of interventions such as exercise or dietary modification on complex metabolic profiles may be useful in guiding CVD prevention strategies in young people.

## Conclusions

This study demonstrates that in an adolescent population, the NMR metabolic profile may present additional targets for lifestyle or pharmaceutical interventions to reduce the harmful effect of adiposity on cardiovascular health, particularly in males. However, our results suggest that in order to have large effects, interventions would require broad approaches to improve whole lipid or lipoprotein profiles and some other small molecules, rather than targeting individual measures. Furthermore, these findings need replication in larger independent samples, analyses to establish causality, and to be explored in adult populations to investigate whether this association is observed with clinical CVD outcomes.

## Data Availability

Code is available from the corresponding author.

## Acknowledgements

We are extremely grateful to all the families who took part in this study, the midwives for their help in recruiting them, and the whole ALSPAC team, which includes interviewers, computer and laboratory technicians, clerical workers, research scientists, volunteers, managers, receptionists and nurses.

This work was carried out using the computational facilities of the Advanced Computing Research Centre -http://www.bris.ac.uk/acrc/ and the Research Data Storage Facility of the University of Bristol - http://www.bris.ac.uk/acrc/storage/.

## Author contributions

ARC analysed and cleaned the data, interpreted results, wrote and revised the manuscript. DLSF advised on statistical analyses, interpreted the results and wrote and revised the manuscript. AET interpreted the results, wrote and revised the manuscript. DAL interpreted the results and critically reviewed and revised the manuscript. GDS interpreted the results and critically reviewed and revised the manuscript. NC provided data for the project, interpreted the results and critically reviewed and revised the manuscript. ADH provided data for the project, interpreted the results and critically reviewed and revised the manuscript. LDH devised the project, advised on statistical analyses, interpreted the results, and wrote and revised the manuscript.

## Funding

ARC, DLSF, AET, DAL, GDS and LDH all work in a unit supported by the UK Medical Research Council and the University of Bristol (Program codes: MC_UU_00011/1 and MC_UU_00011/6]. ARC is supported by a UK Medical Research Council PhD Studentship (MC_UU_00011/1). DAL, AET and GDS are supported by the National Institute for Health Research (NIHR) Biomedical Research Centre based at University Hospitals Bristol NHS Foundation and the University of Bristol. The views expressed are those of the authors and not necessarily those of the NHS, the NIHR, or the Department of Health. DAL’s contribution is supported by the European Union’s Horizon 2020 research and innovation programme under grant agreement No 733206 (LifeCycle). The UK Medical Research Council and Wellcome (Grant ref: 217065/Z/19/Z) and the University of Bristol provide core support for ALSPAC. NMR metabolomics data was funded by the MRC (Grant ref: MC_UU_12013/1) AH received support from the Wellcome Trust (086676/7/08/Z) and the British Heart Foundation (PG/06/145 & CS/15/6/31468) for cardiovascular measures in ALSPAC and works in a unit that receives support from the UK Medical Research Council (Program code: MC_UU_12019/1). LDH is supported by a Career Development Award fellowship from the UK Medical Research Council (MR/M020894/1). No funding body has influenced data collection, analysis or its interpretations. This publication is the work of the authors, who serve as the guarantors for the contents of this paper.

## Disclosures

DAL has received support from several national and international government and charitable funders, as well as Medtronic Ltd and Roche Diagnostics in the last 10-years, for work unrelated to that presented here.

All other authors have no conflicts of interest to declare

Supplemental material

## SUPPLEMENTAL MATERIAL

### EXPANDED METHODS

#### Adiposity measures

Adiposity was measured at age 11. Waist circumference was measured to the nearest millimetre using the Harpenden anthropometric tape. Whole body DXA scans were carried out using a Lunar prodigy narrow fan beam densitometer and used to estimate total-body-less-head fat mass.

#### Cardiac structure measures

Cardiac structure was assessed by echocardiography in a quasi-random subset of participants in ALSPAC at the age 17 clinic. The measures used in this analysis were LVMI, LAI, LVIDD and RWT. Echocardiography was performed using a HDI 5000 ultrasound machine (Philips). All measurements were made according to the American Society of Echocardiography guidelines, where the validated equations were used to calculate LVMI and RWT (23). Average measures of LAI and LVIDD were calculated as the mean of three measurements taken.

#### Covariate measurements

During pregnancy, mothers of ALSPAC children were required to fill in a number of questionnaires answer questions on their age at delivery, the number of pregnancies they have had, their highest educational qualification (less than O-level, O-level, A-level or degree and above), their smoking status (ever versus never), their weight and height before pregnancy (including certainty) and household social class (based on parental occupation, education, type of neighbourhood and use of car).

Offspring covariables included sex (reported at baseline clinics) and birthweight (from birth records, obstetric data and clinic measurements). Adolescent variables considered as confounders were smoking, diet and physical activity. Smoking was self-reported by individuals at age 15 clinics. Participants were defined as a smoker if they had smoked in at least the 30 days prior to attending clinics. Although food frequency questionnaires have been carried out in ALSPAC, these were for a small subset, so although a slightly crude measure of diet fruit and vegetable intake were used to approximate healthy diets. Fruit intake was dichotomised to less than once per day or at least once per day. Vegetable intake was dichotomised to three times or less per week or at least four times per week. Physical activity was defined according to whether the individual takes part in sport with friends as reported by the individual in Focus at 15 clinics.

Age at peak height velocity was considered as a covariate in sensitivity analyses. This was estimated using Superimposition by Translation and Rotation (SITAR) mixed effects growth curve analysis. Repeated measures of height from trained fieldworkers at assessment clinics between the ages of 5 and 20, with at least one measurement for all of ages 5-<10, 10 to <15 and 15 to 20 years were used. For a full description of the methods used see *Frysz* et al (32).

## ADDITIONAL TABLES

**Supplementary Table 1:**
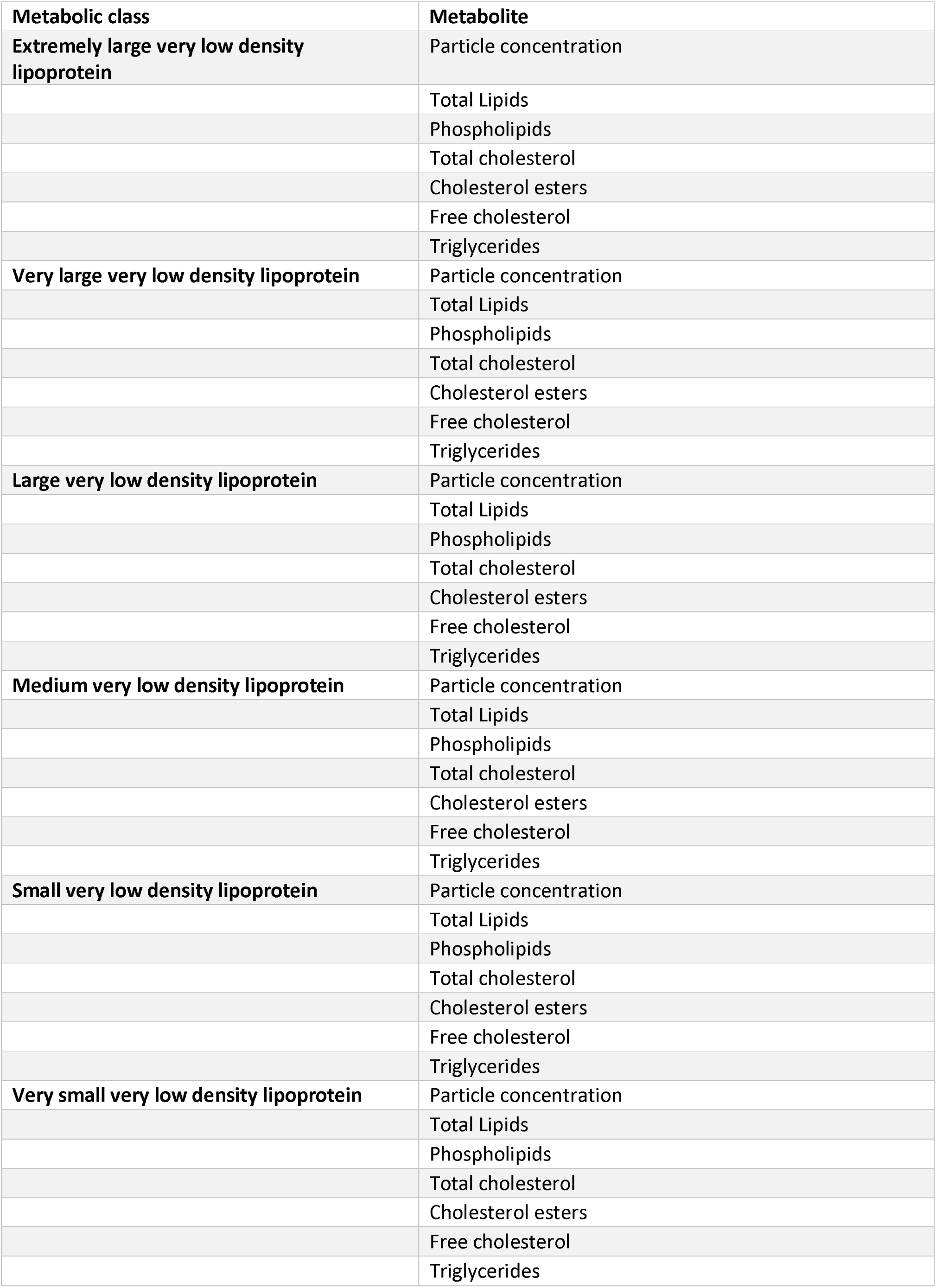
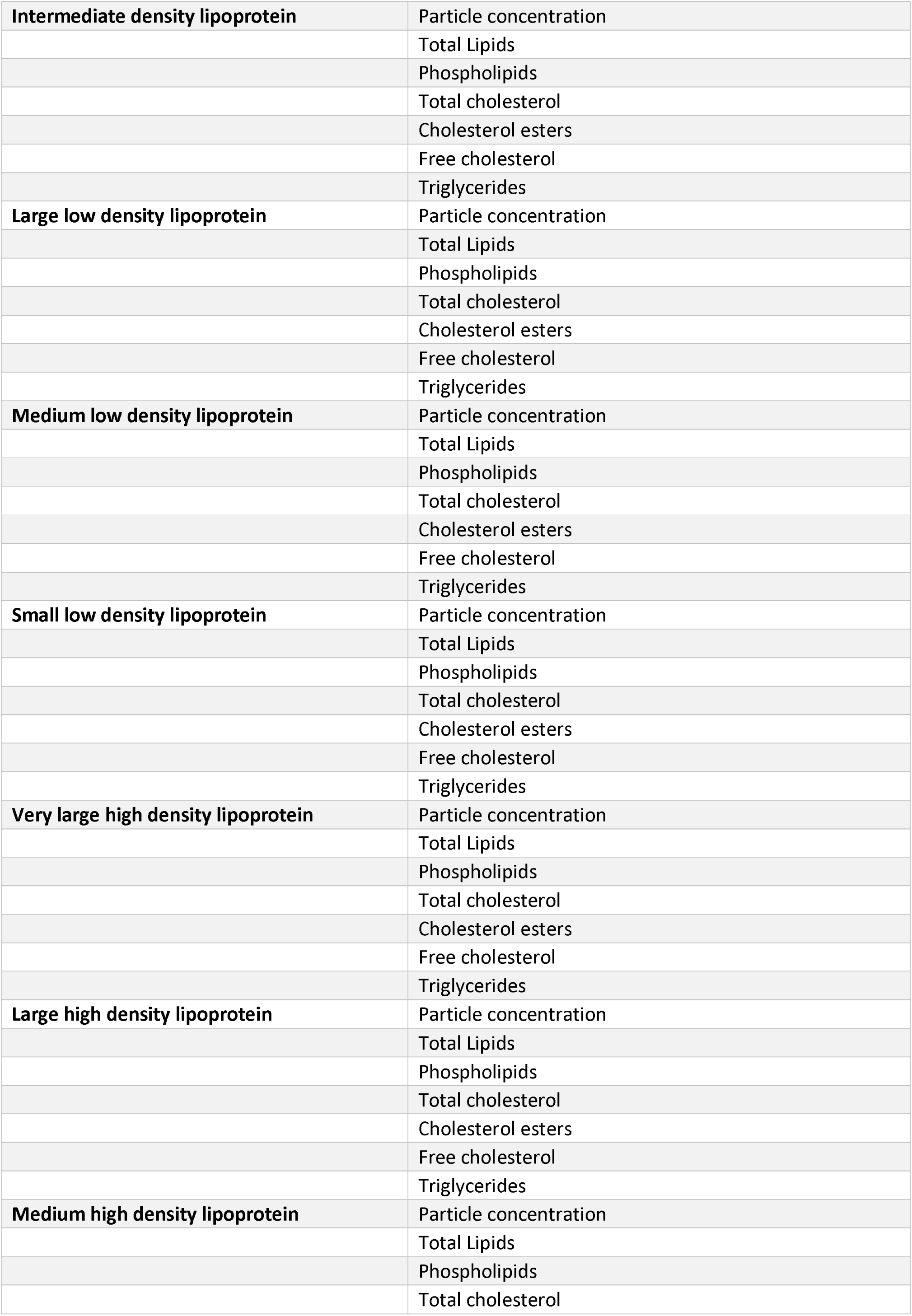
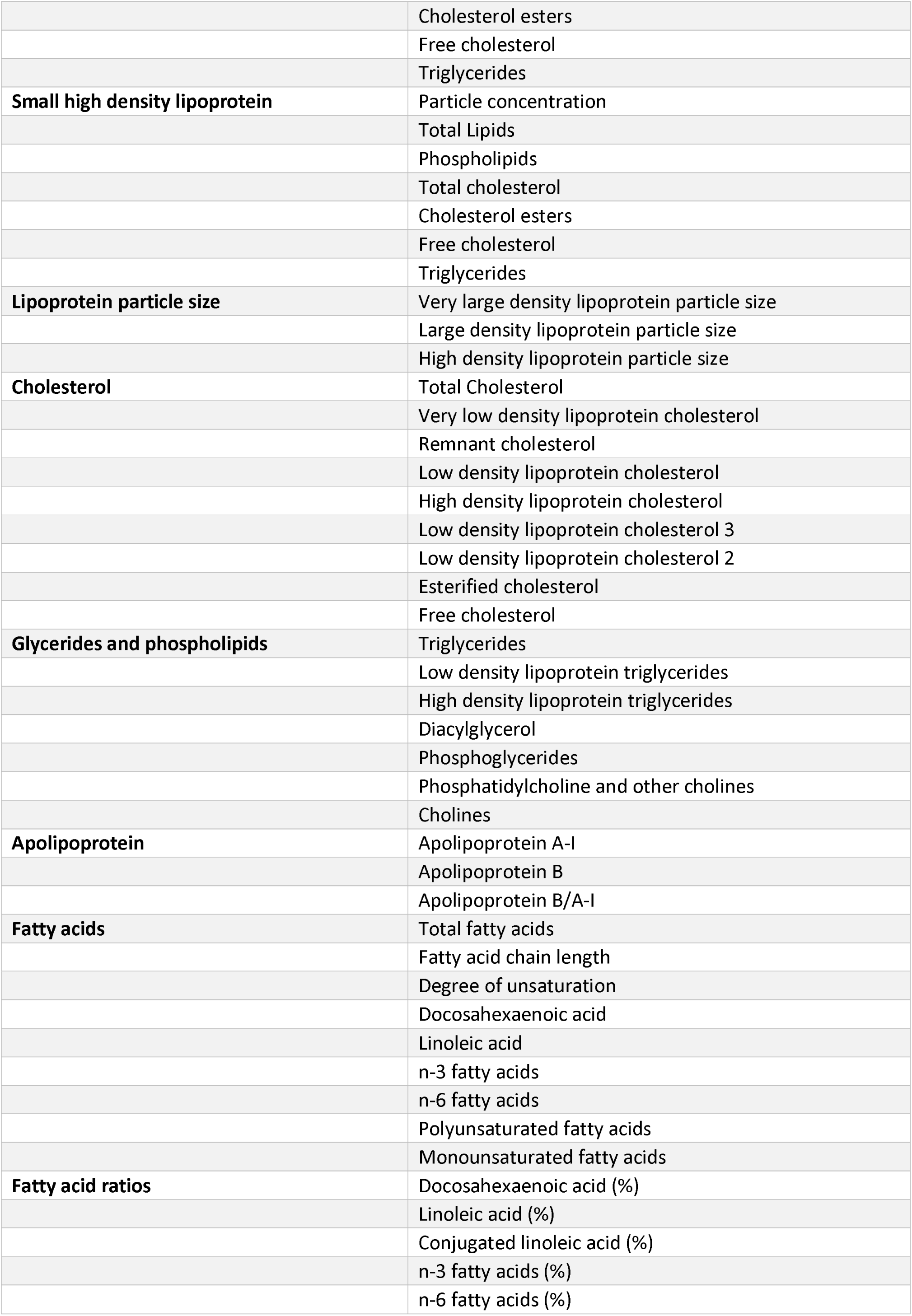
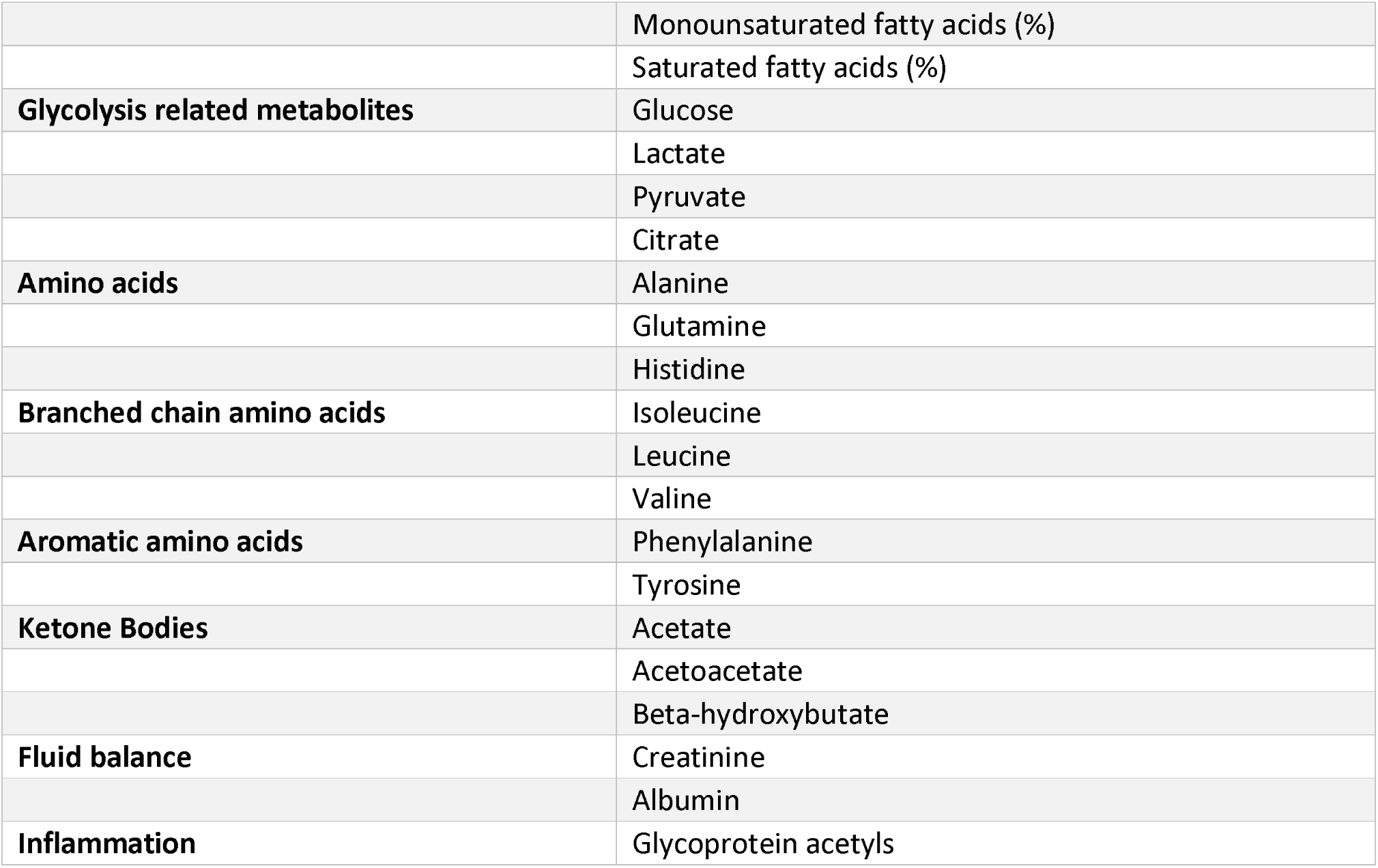
List of metabolites used in analysis.

**Supplementary Table 2:**
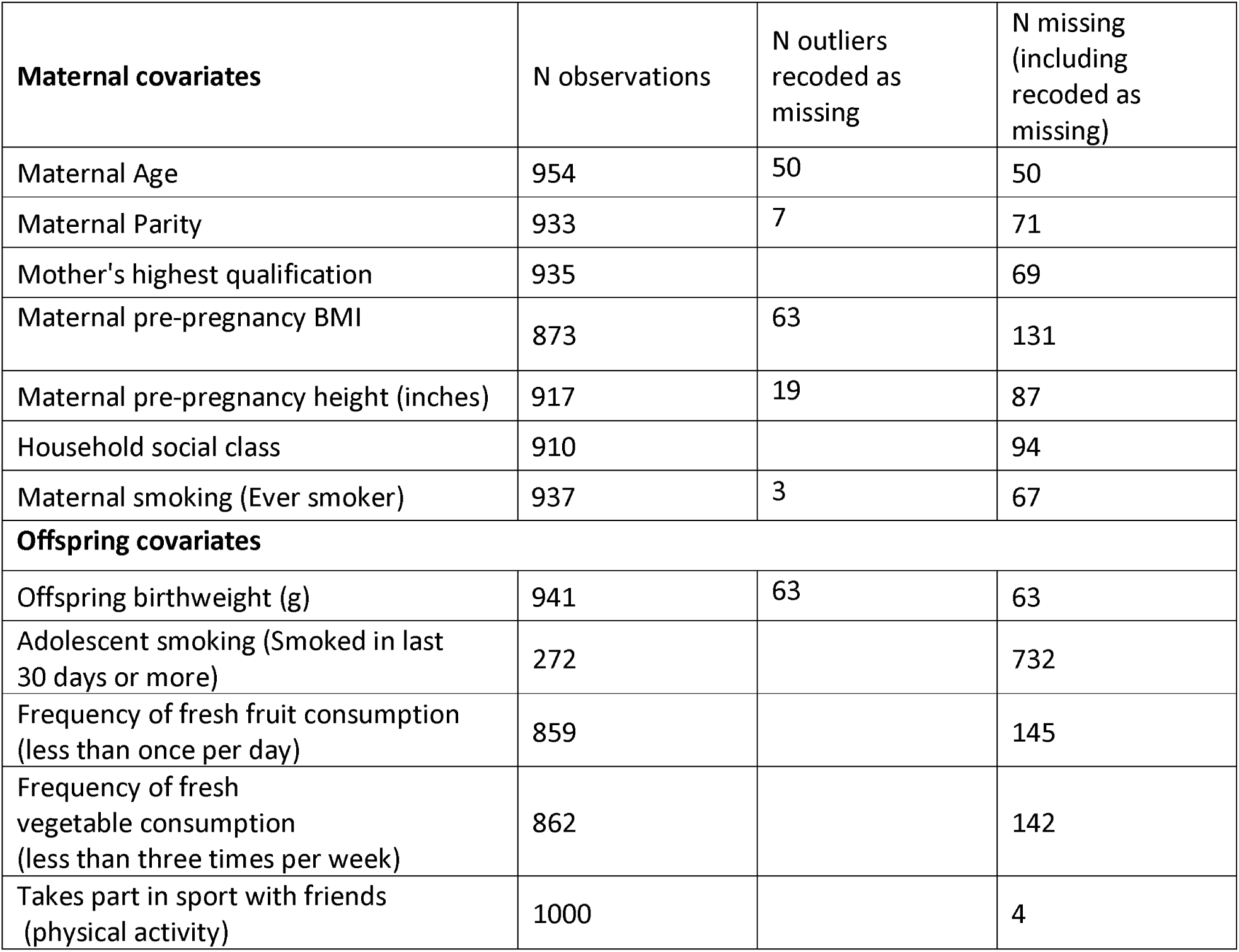
Table of missing data in covariables used in imputations models.

**Supplementary Table 3:** Imputed study characteristics, compared with the non-imputed eligible sample and full ALSPAC cohort.

|  | Imputed Sample (analysis sample)<br>Mean (SD) or proportion (SE) |  |  |  | Eligible Sample<br>Mean (SD) or proportion (SE) |  |  |  | ALSPAC All<br>Mean (SD) or proportion (SE) |  |  |  |
| --- | --- | --- | --- | --- | --- | --- | --- | --- | --- | --- | --- | --- |
|  | N | All | Male | Female | N | All | Male | Female | N | All | Male | Female |
| **Exposures** |  |  |  |  |  |  |  |  |  |  |  |  |
| BMI kg/m<sup>2</sup> | 1004 | 19.07<br>(3.17) | 18.72<br>(3.00) | 19.37<br>(3.29) | 1004 | 19.07 (3.17) | 18.72 (3.00) | 19.37<br>(3.29) | 7106 | 19.11<br>(3.45) | 18.85<br>(3.32) | 19.38<br>(3.56) |
| Waist Circumference (cm) | 1004 | 68.25<br>(8.85) | 68.62<br>(9.03) | 67.93<br>(8.69) | 1004 | 68.25 (8.85) | 68.62 (9.03) | 67.93<br>(8.69) | 7109 | 68.40<br>(9.51) | 68.71<br>(9.73) | 68.15<br>(9.30) |
| Total body fat mass (g) | 1004 | 15217.25<br>(8397.65) | 10889.04<br>(7246.14) | 18981.27<br>(7469.77) | 1004 | 15217.25<br>(8397.65) | 10889.04<br>(7246.14) | 18981.27<br>(7469.77) | 5150 | 15378.78<br>(9220.42) | 11431.67<br>(8458.80) | 19065.85<br>(8369.86) |
| **Outcomes** |  |  |  |  |  |  |  |  |  |  |  |  |
| LVMl g/m<sup>2.7</sup> | 1004 | 28.00<br>(5.87) | 29.92<br>(5.95) | 26.32<br>(5.27) | 1004 | 28.00 (5.87) | 29.92 (5.95) | 26.32<br>(5.27) | 2047 | 27.61<br>(5.98) | 29.43<br>(6.28) | 26.12<br>(5.28) |
| LAI | 1004 | 0.00<br>(0.19) | -0.01<br>(0.24) | 0.00<br>(0.12) | 1004 | 0.00 (0.19) | -0.01 (0.24) | 0.00<br>(0.12) | 1916 | 0.00<br>(0.19) | -0.01<br>(0.24) | 0.00<br>(0.15) |
| RWT | 1004 | 0.38<br>(0.06) | 0.39<br>(0.06) | 0.37<br>(0.06) | 1004 | 0.38 (0.06) | 0.39 (0.06) | 0.37<br>(0.06) | 2056 | 0.38<br>(0.06) | 0.38<br>(0.06) | 0.38<br>(0.06) |
| LVIDD Average (cm) | 1004 | 4.53<br>(0.46) | 4.73<br>(0.49) | 4.35<br>(0.36) | 1004 | 4.53 (0.46) | 4.73 (0.49) | 4.35<br>(0.36) | 2118 | 4.50<br>(0.44) | 4.74<br>(0.42) | 4.74<br>(0.42) |
| **Covariates (offspring)** |  |  |  |  |  |  |  |  |  |  |  |  |
| Sex (% Male) | 1004 |  |  |  | 1004 | 0.30 (0.03) |  |  | 14 834 | 0.38 (0.01) |  |  |
| Offspring birthweight (g) | 1004 | 3463.90<br>(525.00) | 3549.90<br>(558.76) | 3389.12<br>(481.91) | 941 | 3465.45<br>(514.14) | 3555.60<br>(538.58) | 3386.61<br>(478.56) | 13 883 | 3381.51<br>(580.57) | 3443.46<br>(595.93) | 3339.49<br>(536.85) |
| Adolescent smoking (Smoked in last 30 days or more) | 1004 | 0.54<br>(0.03) | 0.54<br>(0.06) | 0.54<br>(0.03) | 272 | 0.55 (0.03) | 0.58 (0.06) | 0.54<br>(0.04) | 1719 | 0.55<br>(0.01) | 0.57<br>(0.02) | 0.54<br>(0.02) |
| Frequency of fresh fruit consumption (less than once per day) | 1004 | 0.83<br>(0.01) | 0.85<br>(0.02) | 0.82<br>(0.02) | 859 | 0.85 (0.02) | 0.86 (0.14) | 0.85<br>(0.03) | 8373 | 0.85<br>(0.01) | 0.85<br>(0.17) | 0.85<br>(0.01) |
| Frequency of fresh vegetable consumption (less than three times per week) | 1004 | 0.72 (0.01) | 0.71 (0.02) | 0.72 (0.02) | 862 | 0.77 (0.03) | 0.71 (0.06) | 0.80 (0.03) | 8400 | 0.74 (0.01) | 0.80 (0.02) | 0.75 (0.02) |
| Takes part in sport with friends (physical activity) | 1004 | 0.64 (0.02) | 0.75 (0.02) | 0.55 (0.02) | 1000 | 0.65 (0.03) | 0.77 (0.05) | 0.60 (0.04) | 7087 | 0.65 (0.01) | 0.77 (0.23) | 0.57 (0.02) |
| Covariates (maternal) |  |  |  |  |  |  |  |  |  |  |  |  |
| Maternal Age | 1004 | 29.50 (4.45) | 29.63 (4.30) | 29.40 (4.58) | 954 | 29.50 (4.44) | 29.63 (4.28) | 29.39 (4.57) | 14 062 | 27.99 (4.97) | 28.08 (5.01) | 27.87 (4.93) |
| Maternal Parity | 1004 | 0.70 (0.83) | 0.68 (0.83) | 0.72 (0.83) | 933 | 0.70 (0.82) | 0.68 (0.83) | 0.72 (0.82) | 13 111 | 0.84 (1.00) | 0.86 (1.03) | 0.83 (0.97) |
| Mother's highest qualification | 1004 | 2.56 (0.99) | 2.61 (0.98) | 2.50 (1.00) | 935 | 2.56 (0.99) | 2.63 (0.97) | 2.50 (1.00) | 12 323 | 2.18 (1.00) | 2.56 (1.07) | 2.19 (1.01) |
| Maternal pre-pregnancy BMI | 1004 | 22.95 (3.57) | 22.98 (3.43) | 22.92 (3.70) | 873 | 22.93 (3.54) | 22.97 (3.38) | 22.89 (3.68) | 11 670 | 22.92 (3.83) | 22.97 (3.84) | 22.89 (3.82) |
| Maternal pre-pregnancy height (inches) | 1004 | 64.68 (2.68) | 64.78 (2.82) | 64.60 (2.56) | 917 | 64.68 (2.56) | 64.78 (2.58) | 64.59 (2.54) | 12 370 | 64.56 (2.65) | 64.57 (2.67) | 64.53 (2.62) |
| Household social class | 1004 | 2.28 (1.01) | 2.22 (1.02) | 2.32 (0.99) | 910 | 2.27 (1.00) | 2.21 (1.02) | 2.31 (0.98) | 11 416 | 2.57 (1.07) | 2.56 (1.07) | 2.57 (1.07) |
| Maternal smoking (Ever smoker) | 1004 | 0.38 (0.02) | 0.36 (0.02) | 0.40 (0.02) | 937 | 0.49 (0.03) | 0.48 (0.06) | 0.49 (0.04) | 13 236 | 0.51 (0.01) | 0.52 (0.02) | 0.51 (0.02) |

**Supplementary Table 4:** Total effects between adiposity and cardiac structure, excluding and including an interaction parameter for sex (complete case analysis)

| Exposure | Outcome | Beta with no interaction parameter (95% CI) | Beta with interaction parameter (95% CI) | P value for interaction |
| --- | --- | --- | --- | --- |
| **BMI** | LVMl | 0.799 (0.567, 1.032) | 0.688 (0.266, 1.11) | 0.514 |
|  | LAI | -0.009 (-0.015, -0.003) | -0.015 (-0.026, -0.004) | 0.216 |
|  | RWT | 0.001 (-0.001, 0.004) | 0.002 (-0.003, 0.006) | 0.974 |
|  | LVIDD | 0.035 (0.017, 0.053) | 0.036 (0.003, 0.068) | 0.970 |
| **Waist Circumference** | LVMl | 0.242 (0.153, 0.33) | 0.191 (0.045, 0.336) | 0.361 |
|  | LAI | -0.003 (-0.006, -0.001) | -0.005 (-0.008, -0.001) | 0.334 |
|  | RWT | 0.001 (0, 0.002) | 0.001 (-0.001, 0.002) | 0.733 |
|  | LVIDD | 0.011 (0.005, 0.018) | 0.011 (0, 0.022) | 0.945 |
| **DXA** | LVMl | 2.53E-04<br>(1.53E-04, 3.54E-04) | 1.42E-04<br>(-0.000024, 3.07E-04) | 0.080 |
|  | LAI | -3.1E-06<br>(-5.6E-06, -5.5E-07) | -4E-06<br>(-8.2E-06, 2.11E-07) | 0.578 |
|  | RWT | 6.88E-07<br>(-4.3E-07, 1.8E-06) | 7.33E-07<br>(-1.1E-06, 2.58E-06) | 0.949 |
|  | LVIDD | 1.15E-05<br>(4.06E-06, 1.89E-05) | 5.33E-06<br>(-6.9E-06, 1.76E-05) | 0.193 |

**Supplementary Table 5:** **The proportion mediated by standard cardiovascular risk factors alone, metabolites considered jointly as principal components and standard cardiovascular risk factors in addition to metabolite principle components on the association between BMI and left ventricular mass, adjusting for peak height velocity as a covariate**

| Mediator | Female |  | Male |  |
| --- | --- | --- | --- | --- |
|  | Indirect effect | Proportion mediated | Indirect effect | Proportion mediated |
| **Established risk factors** | 0.02 (-0.02, 0.06) | 2.77 (-2.31, 7.86) | 0.02 (0.00, 0.05) | 2.77 (0.28, 5.26) |
| **Metabolites only** | 0.03 (-0.04, 0.09) | 3.39 (-13.72, 20.5) | 0.03 (-0.02, 0.08) | 3.8 (-8.65, 16.26) |
| **Established risk factors plus metabolites** | 0.02 (0.00, 0.05) | 2.77 (0.28, 5.26) | 0.08 (-0.14, 0.31) | 10.68 (-0.86, 22.23) |

**Supplementary Table 6.** Total effects between adiposity and cardiac structure for complete case analysis and multiply imputed data.

| Exposure | Outcome | Female |  | Male |  |
| --- | --- | --- | --- | --- | --- |
|  |  | Complete Case (N = 184) | Multiply Imputed (N = 536) | Complete Case (N = 55) | Multiply Imputed (N = 437) |
| **BMI** | LVMl | 0.831 (0.559, 1.102) | 0.661 (0.529, 0.793) | 0.620 (0.167, 1.072) | 0.701 (0.525, 0.877) |
|  | LAI | -0.007 (-0.013, -0.001) | -0.002 (-0.006, 0.001) | -0.017 (-0.034, -0.001) | -0.006 (-0.016, 0.003) |
|  | RWT | 0.001 (-0.002, 0.005) | 0.001 (-0.0001, 0.003) | 0.001 (-0.004, 0.005) | 0.002 (2.86E-05, 0.004) |
|  | LVIDD | 0.035 (0.012, 0.057) | 0.027 (0.017, 0.036) | 0.036 (0.005, 0.067) | 0.012 (0.027, 0.042) |
| **Waist Circumference** | LVMl | 0.251 (0.139, 0.363) | 0.192 (0.14, 0.243) | 0.167 (0.013, 0.322) | 0.188 (0.128, 0.248) |
|  | LAI | -0.003 (-0.005, 0.0004) | -0.001 (-0.002, 0) | -0.006 (-0.011, -0.0002) | -0.003 (-0.006, 0) |
|  | RWT | 0.001 (-0.001, 0.002) | 0.001 (0, 0.001) | 0.0003 (-0.001, 0.002) | 0.001 (0, 0.001) |
|  | LVIDD | 0.011 (0.002, 0.020) | 0.009 (0.006, 0.013) | 0.011 (0.0002, 0.021) | 0.009 (0.004, 0.014) |
| **DXA** | LVMl | 3.18E-04 (1.95E-04, 4.41E-04) | 2.78E-04 (2.18E-04, 3.39E-04) | 1.36E-04 (-3.50E-05, 3.06E-04) | 2.09E-04 (1.35E-04, 2.84E-04) |
|  | LAI | -2.75E-06 (-5.28E-06, -2.16E-07) | -7.30E-07 (-2.19E-06, 7.30E-07) | 3.78E-06 (9.75E-06, 2.18E-06) | -1.60E-06 (-4.95E-06, 1.75E-06) |
|  | RWT | 6.74E-07 (9.17E-07, 2.27E-06) | 7.47E-07 (1.93E-08, 1.47E-06) | 5.18E-07 (-1.05E-06, 2.18E-06) | 1.33E-06 (5.27E-07, 2.13E-06) |
|  | LVIDD | 1.55E-05 (5.71E-06, 2.53E-05) | 1.38E-05 (9.64E-06, 1.81E-05) | 5.35E-06 (-6.44E-06, 1.72E-05) | 4.76E-06 (-1.24E-06, 1.08E-05) |

## ADDITIONAL FIGURES

**Supplementary Figure 1:**
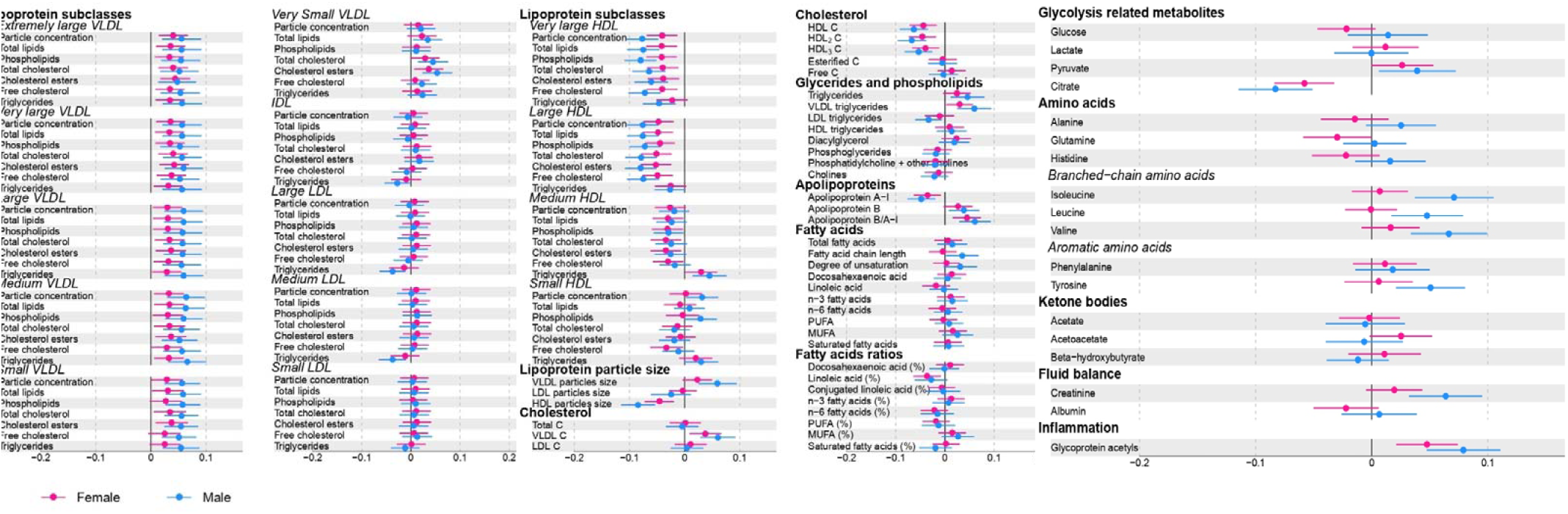
The association between BMI and individual metabolic traits.

**Supplementary Figure 2:**
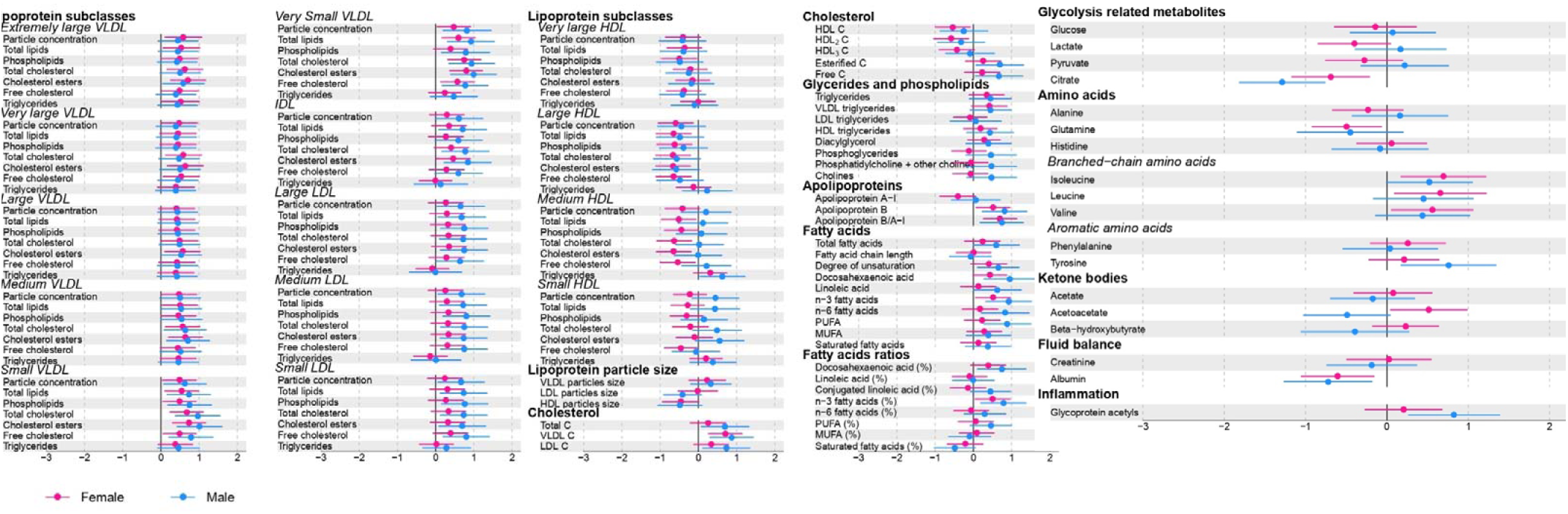
The association between individual metabolic traits and left ventricular mass indexed to height^2.7^.

**Supplementary Figure 3:**
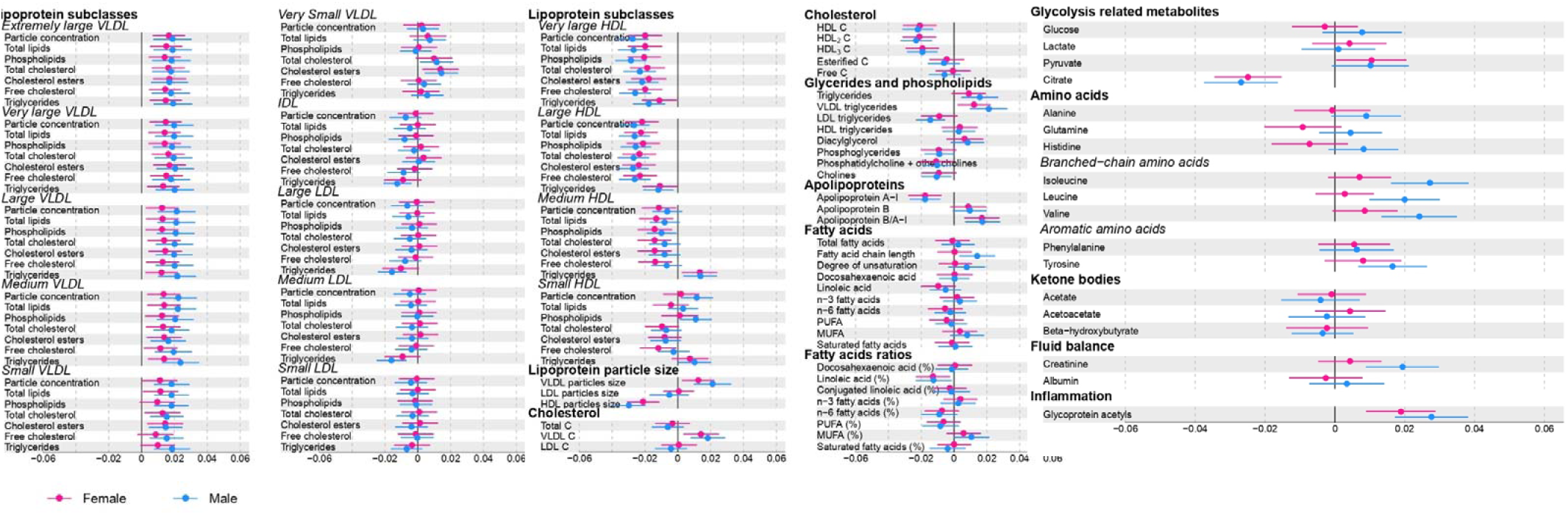
The association between waist circumference and individual metabolic traits.

**Supplementary Figure 4:**
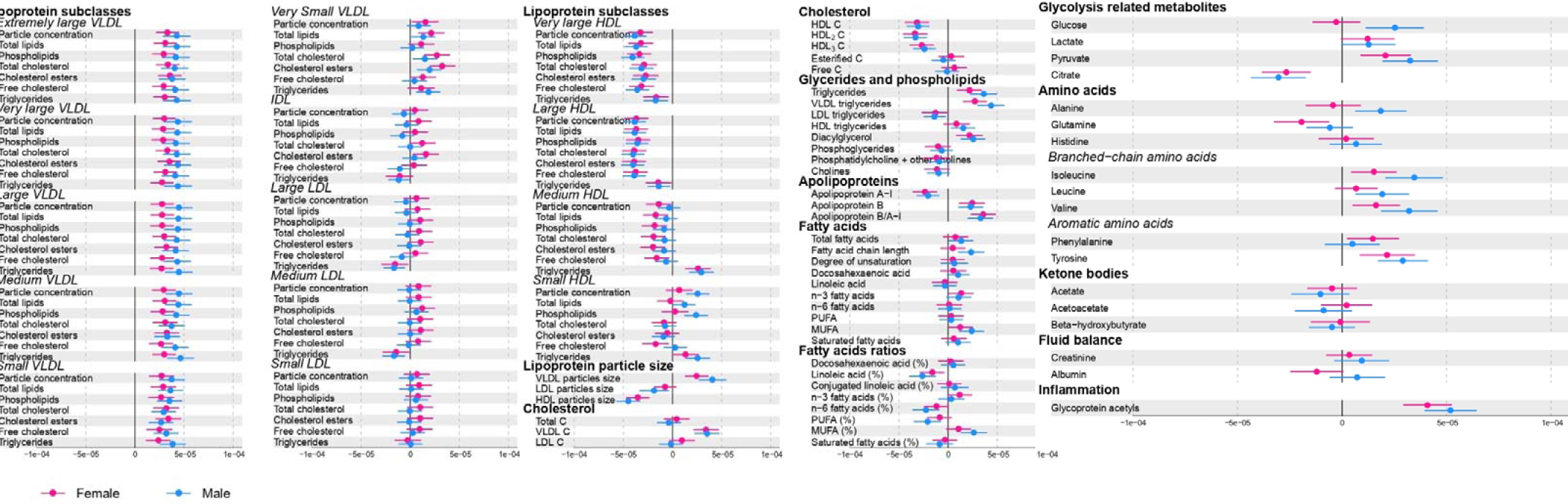
The association between DXA-determined fat mass and individual metabolic traits.

**Supplementary Figure 5:**
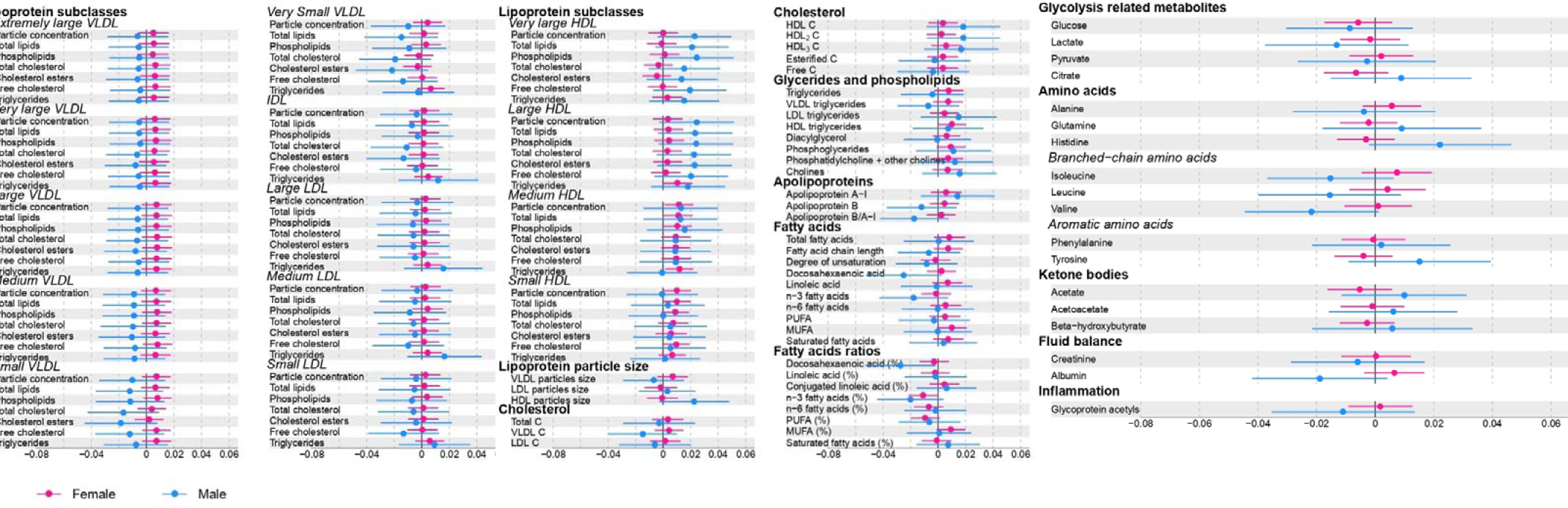
The association between individual metabolic traits and left atrial size indexed to height.

**Supplementary Figure 6:**
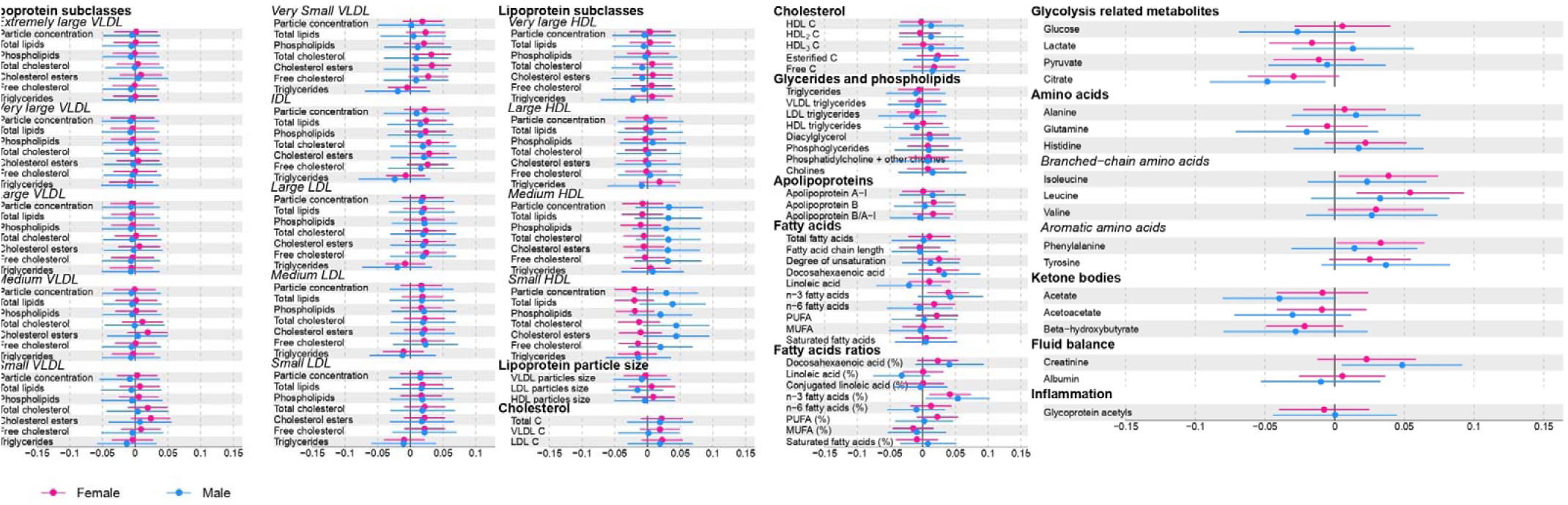
The association between individual metabolic traits and left ventricular internal diameter.

**Supplementary Figure 7:**
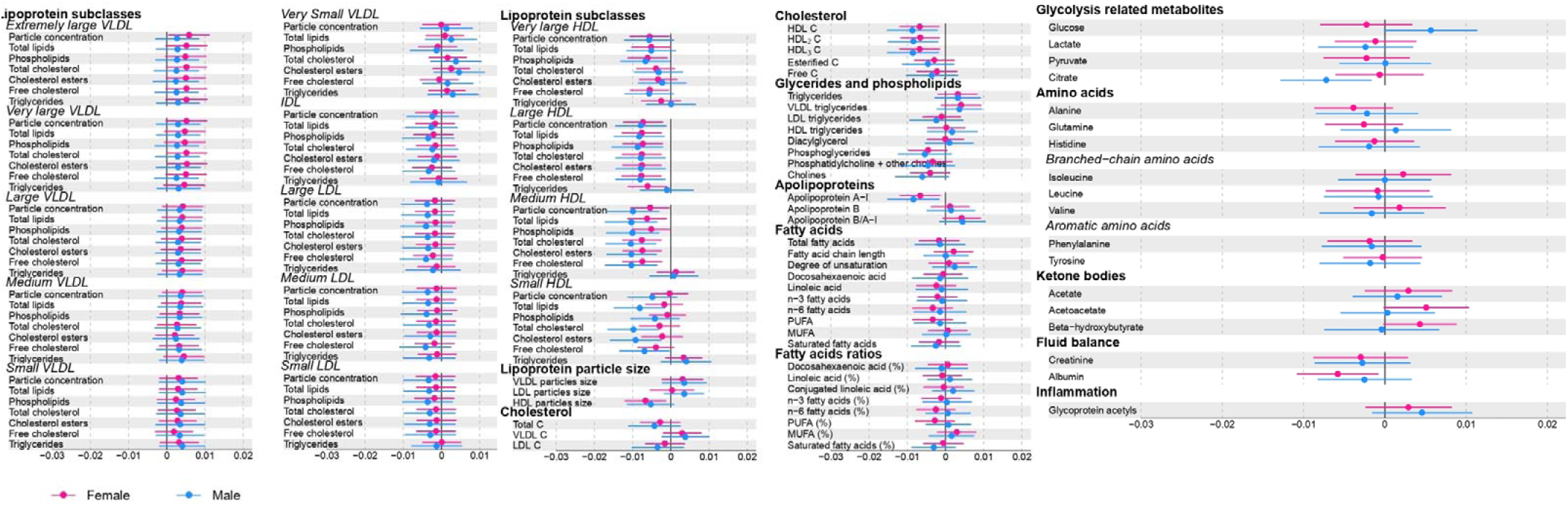
The association between individual metabolic traits and relative wall thickness.

**Supplementary Figure 8:**
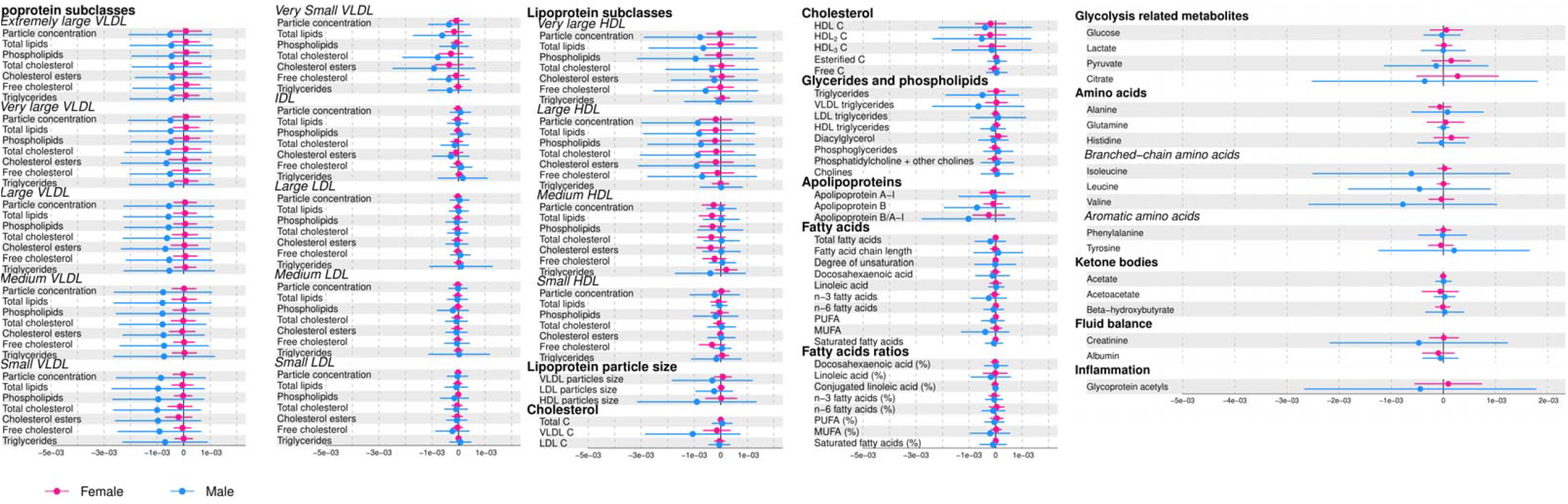
The indirect effect explained by each individual metabolic trait for the association of body mass index and left atrial size indexed to height.

**Supplementary Figure 9:**
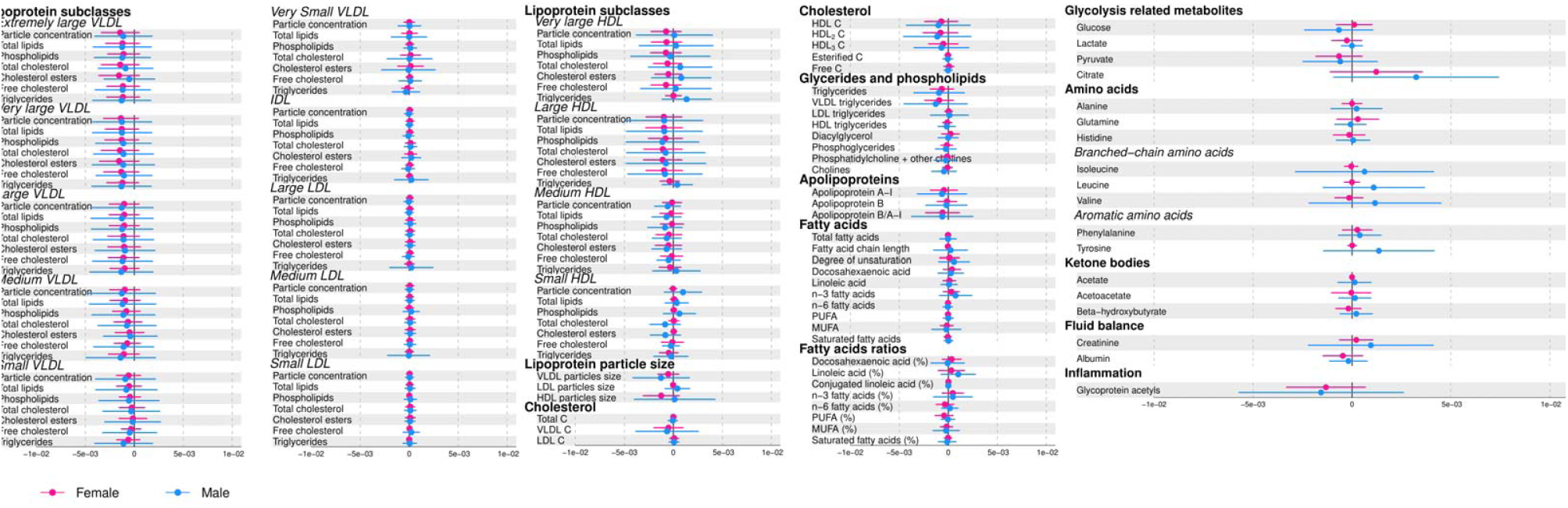
The indirect effect amount explained by each individual metabolic trait for the association of body mass index and left ventricular internal diameter.

**Supplementary Figure 10:**
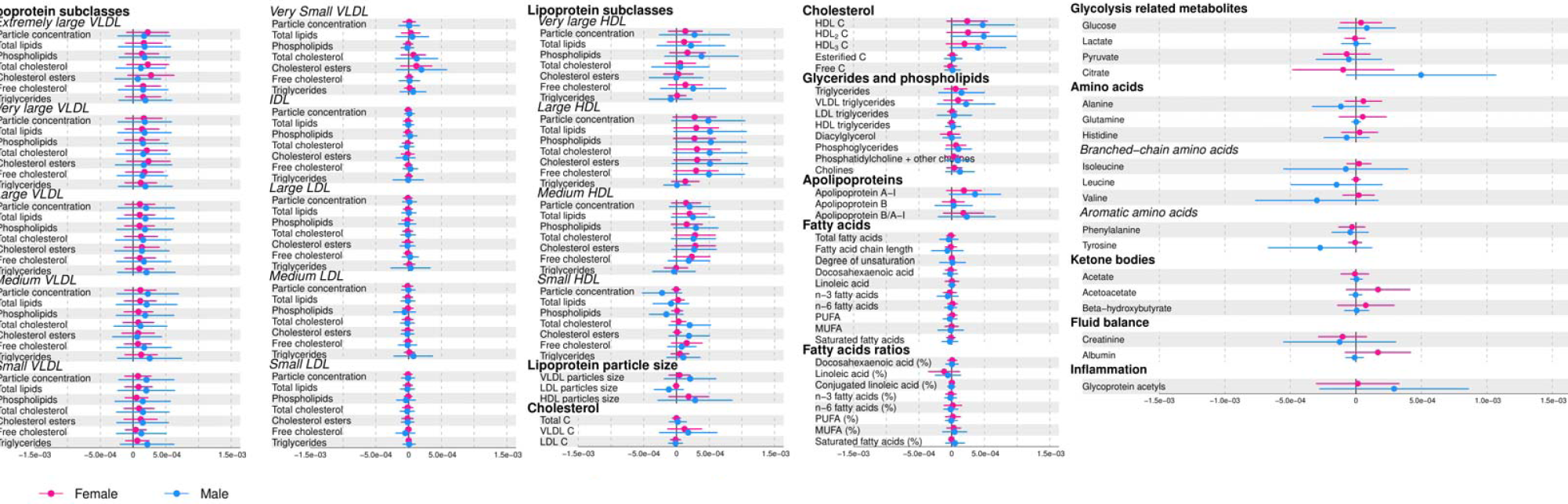
The indirect effect amount explained by each individual metabolic trait for the association of body mass index and relative wall thickness.

**Supplementary Figure 11:**
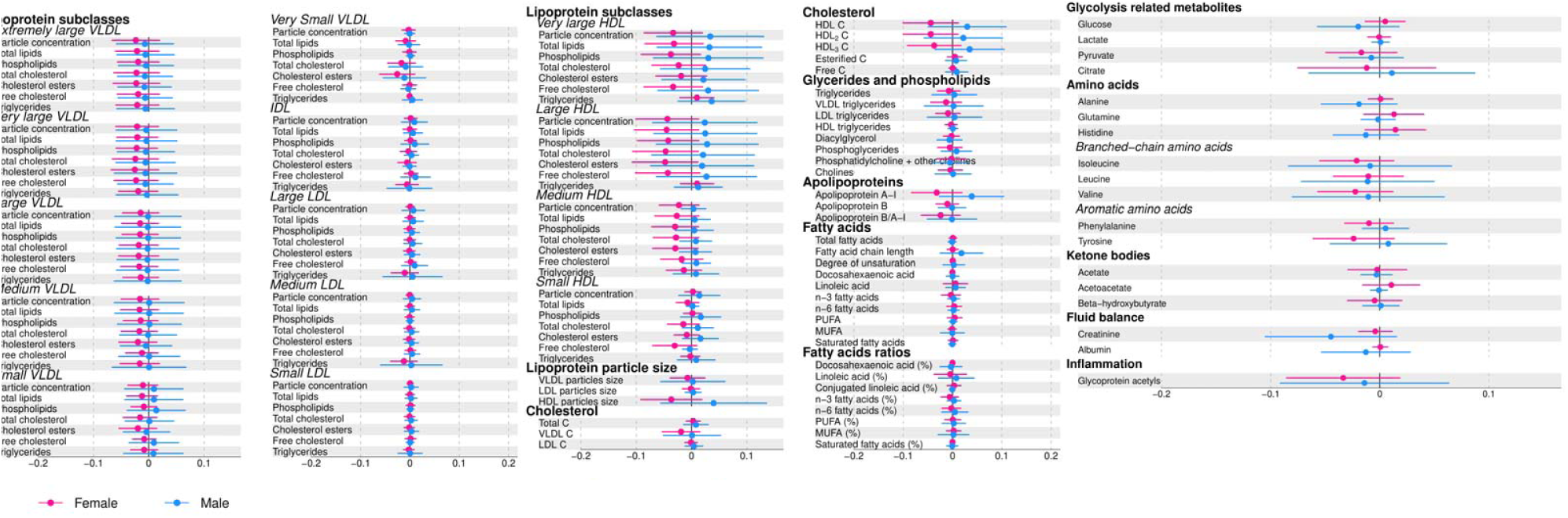
The indirect effect amount explained by each individual metabolic trait for the association of waist circumference and left ventricular mass indexed to height^2.7^.

**Supplementary Figure 12:**
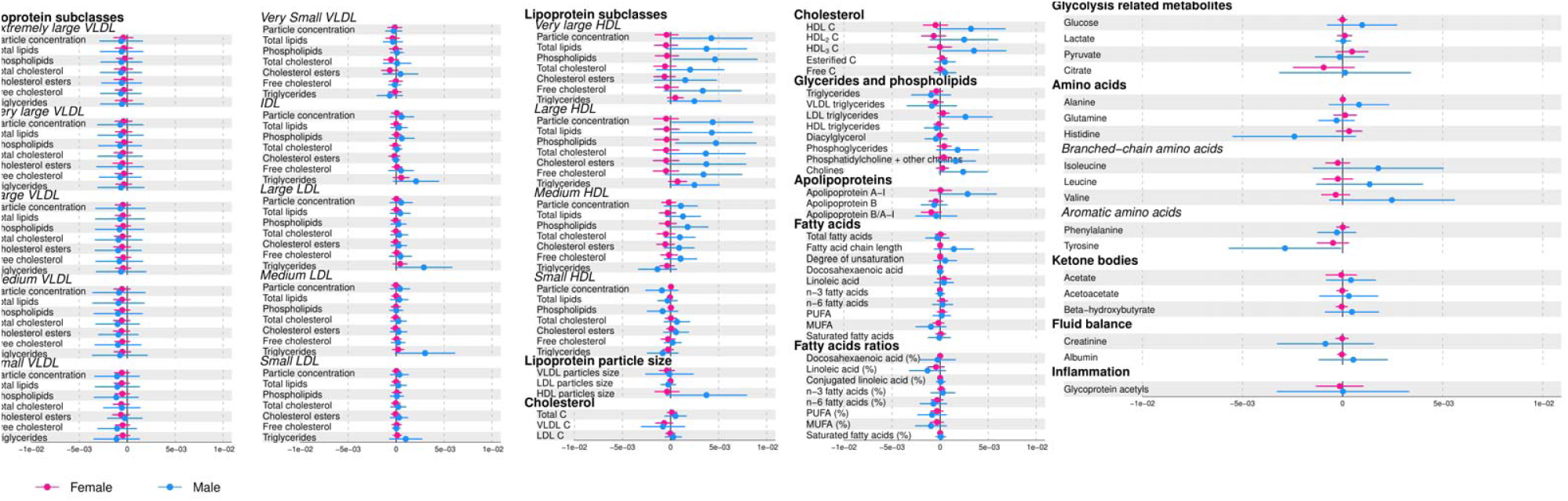
The indirect effect amount explained by each individual metabolic trait for the association of waist circumference and left atrial size indexed to height.

**Supplementary Figure 13:**
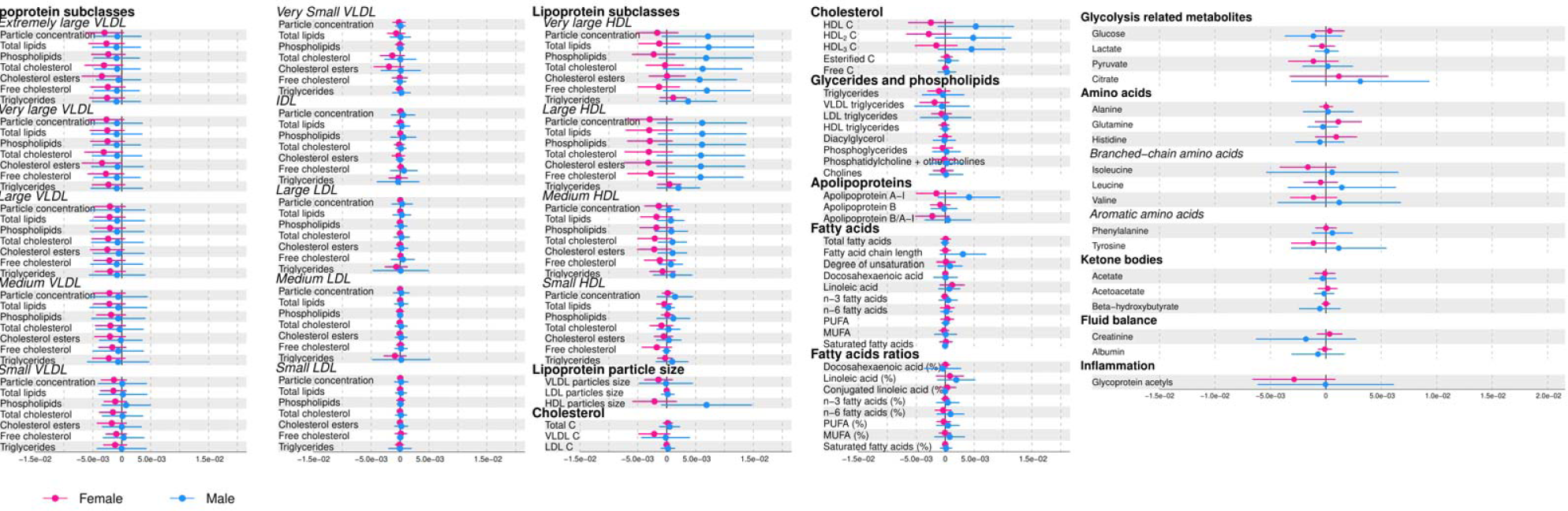
The indirect effect amount explained by each individual metabolic trait for the association of waist circumference and left ventricular internal diameter.

**Supplementary Figure 14:**
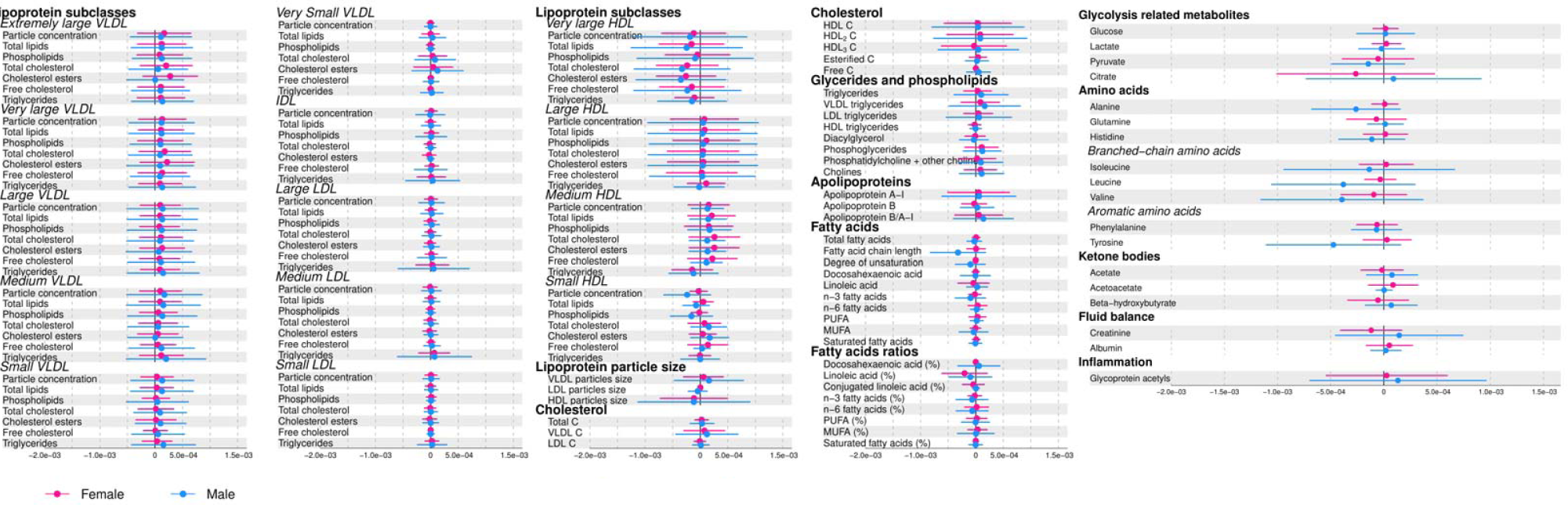
The indirect effect amount explained by each individual metabolic trait for the association of waist circumference and relative wall thickness.

**Supplementary Figure 15:**
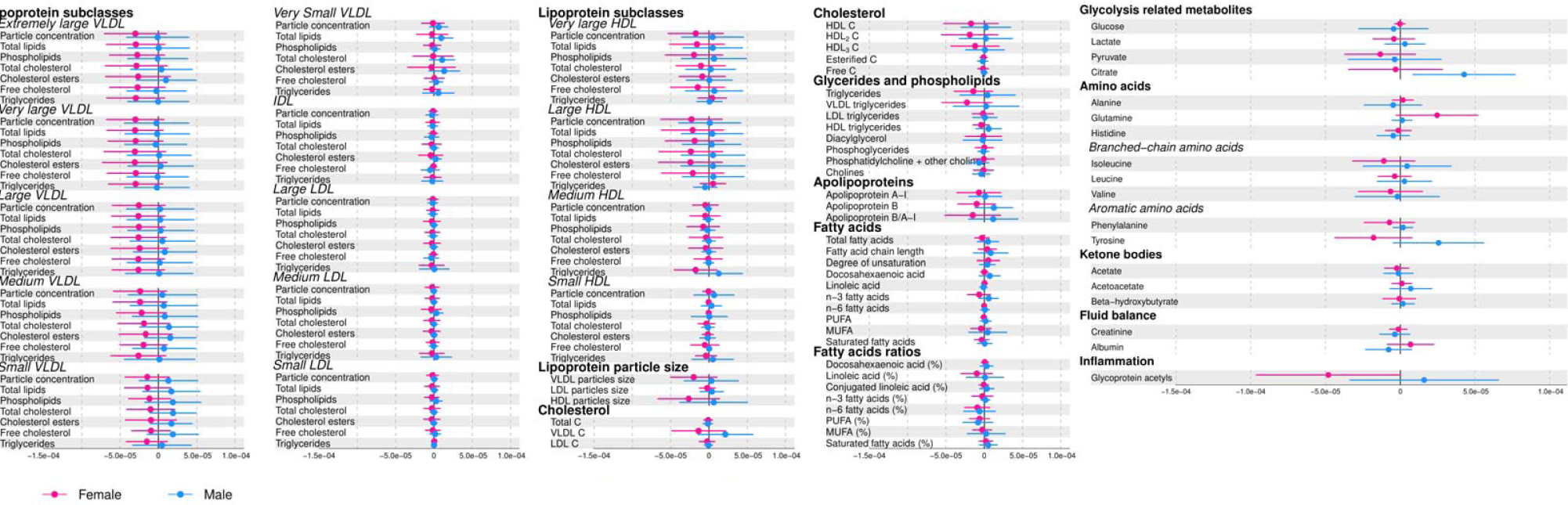
The indirect effect amount explained by each individual metabolic trait for the association of DXA-determined fat mass and left ventricular mass indexed to height^2.7^.

**Supplementary Figure 16:**
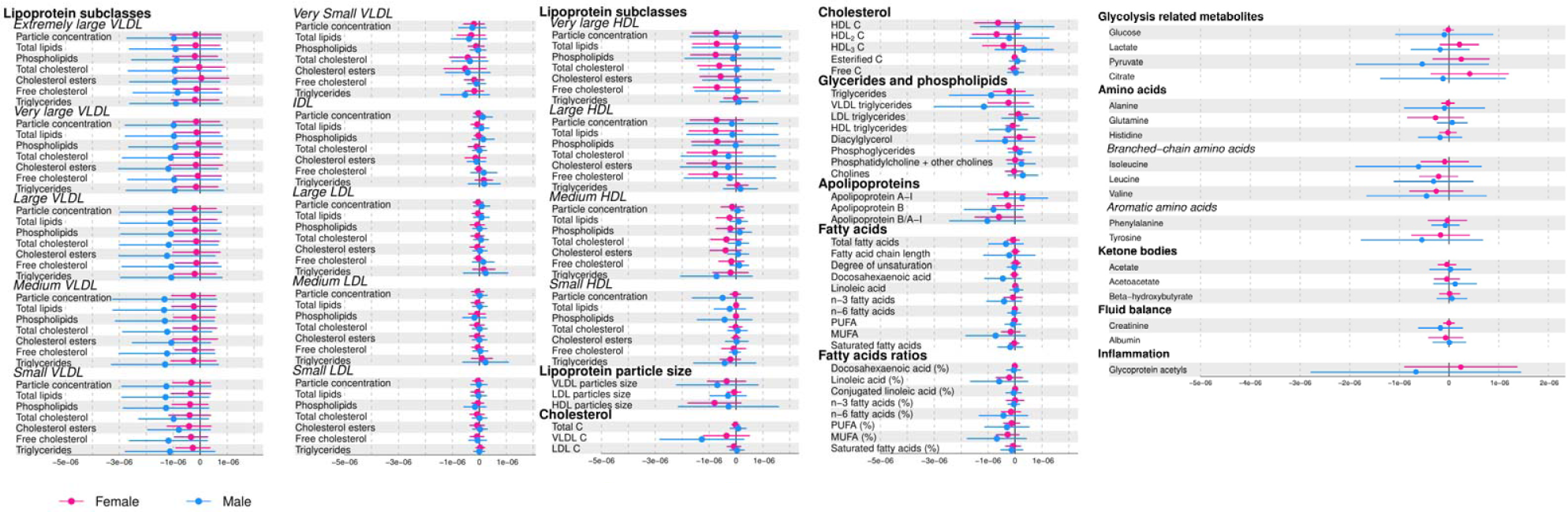
The indirect effect amount explained by each individual metabolic trait for the association of DXA-determined fat mass and left atrial size indexed to height.

**Supplementary Figure 17:**
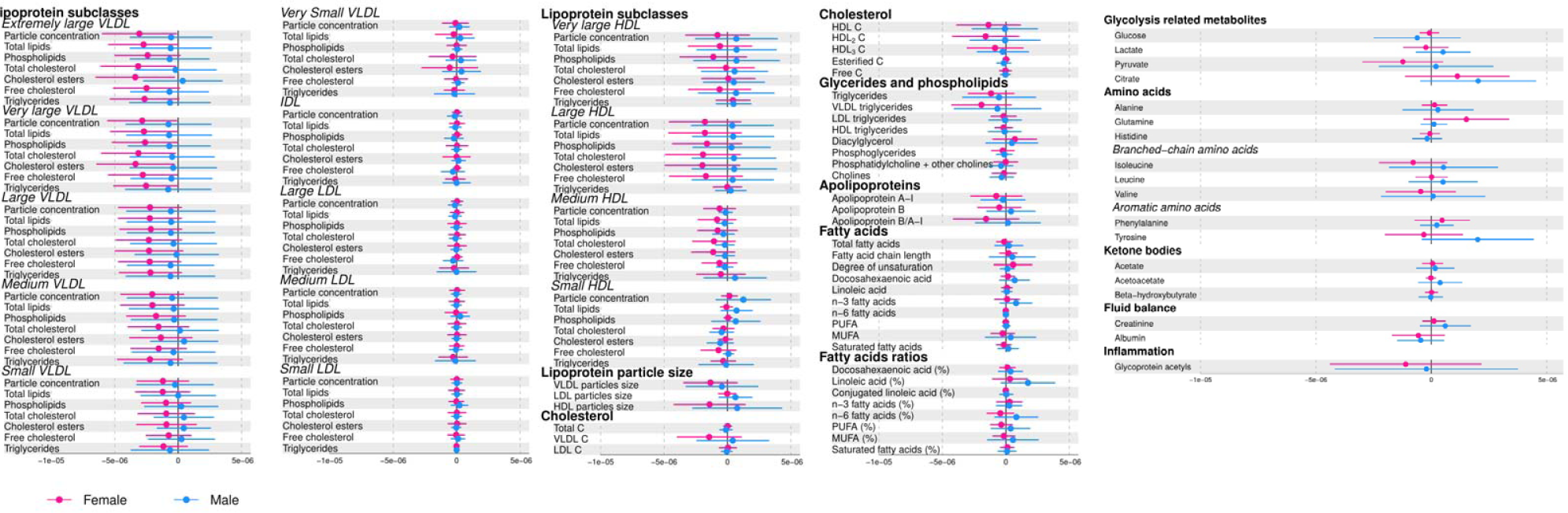
The indirect effect amount explained by each individual metabolic trait for the association of DXA-determined fat mass and left ventricular internal diameter.

**Supplementary Figure 18:**
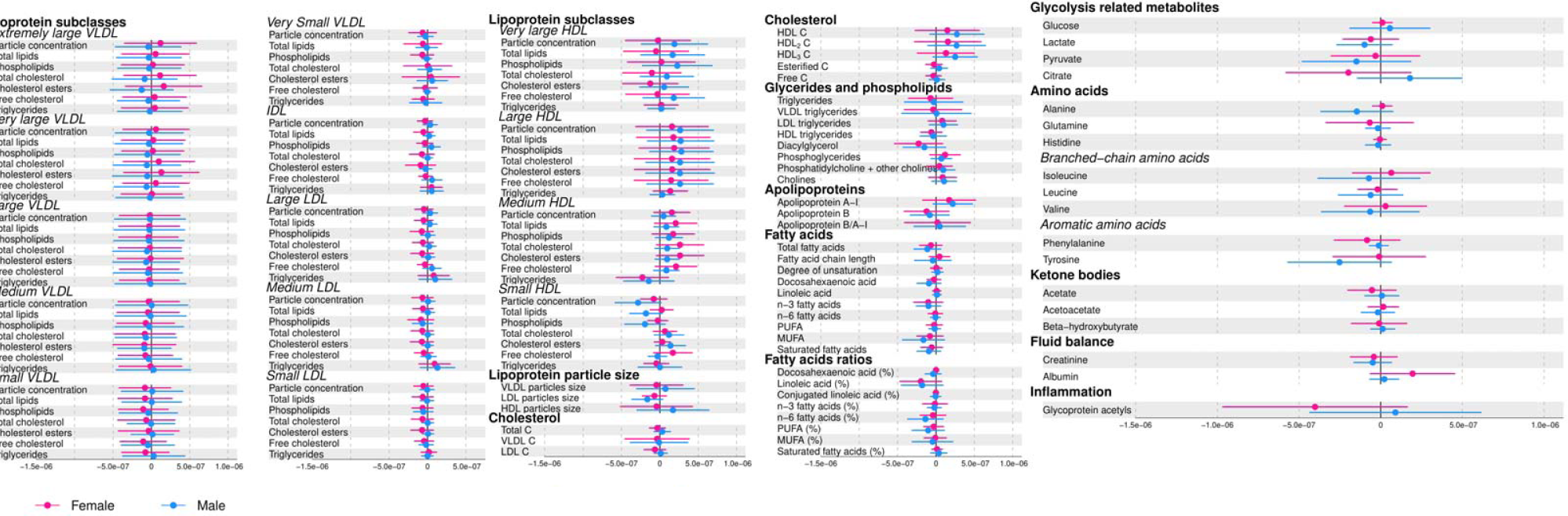
The indirect effect amount explained by each individual metabolic trait for the association of DXA-determined fat mass and relative wall thickness.

**Supplementary Figure 19:**
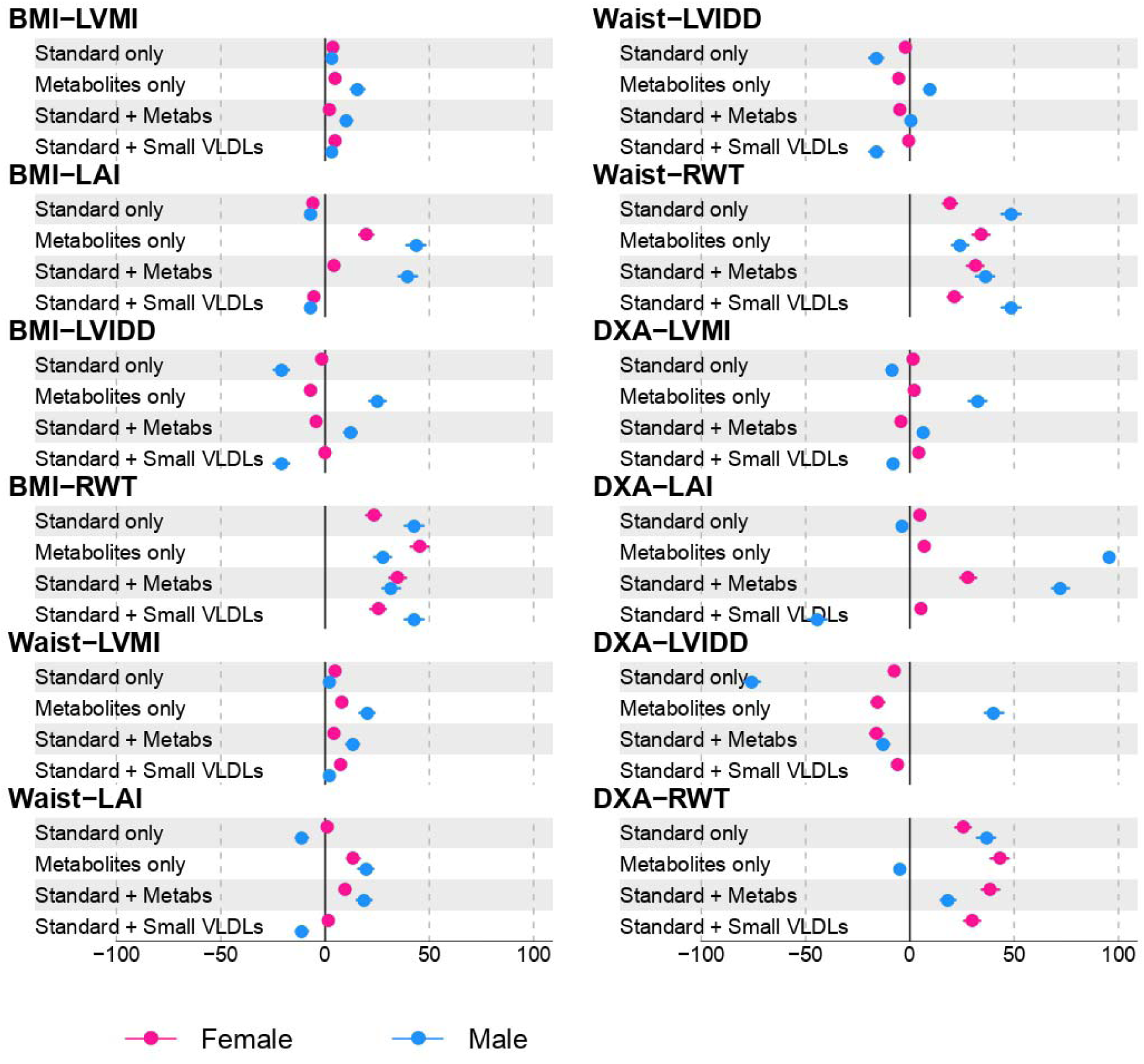
Forest plot showing the proportion mediated by measures of adiposity (Body mass index [BMI], waist circumference [waist] and dual x-ray absorptiometry [DXA]-determined fat mass) with cardiac structure (left atrial size indexed to height [LAI], left ventricular mass indexed to height^2.7^ [LVMI], left ventricular internal diameter [LVIDD] and relative wall thickness [RWT]) measured using electrocardiography. Mediation was considered by i) standard risk factors ii) metabolic principal components (explaining 95% of the variation in the metabolic profile) iii) established risk factors plus metabolic PCs and iv) standard risk factors and small very low-density lipoproteins (VLDLs) Standard mediators: systolic blood pressure, diastolic blood pressure, insulin, low density lipoprotein and glucose. Models adjusted for: Maternal age, Maternal parity, Maternal education, Maternal pre-pregnancy height, Maternal pre-pregnancy BMI, maternal smoking, household social class and adolescent birthweight Note: Models for the effect of standard mediators plus small VLDLS in males for the association between DXA-determined fat mass and LVIDD and DXA-determined fat mass and RWT were out of the bounds of reasonable interpretation.

